# A scoping review on control strategies for *Echinococcus granulosus sensu lato*

**DOI:** 10.1101/2024.08.21.24312335

**Authors:** Tania De la Cruz-Saldana, Javier A. Bustos, Maria P. Requena-Herrera, Nelson Martinez-Merizalde, Lizzie Ortiz-Cam, Ana Lucía Cáceres, Carolina Guzman, Cesar M. Gavidia, Cesar Ugarte-Gil, Ricardo Castillo-Neyra

## Abstract

**Background:** Cystic echinococcosis (CE) is a widespread neglected zoonotic disease caused by *Echinococcus granulosus sensu lato* (EG) with a global burden of control in the billions of dollars. *E. granulosus’* life cycle involves definitive, intermediate, and humans as dead-end hosts. Echinococcosis control programs use strategies that focus on any of these hosts. We aimed to provide a comprehensive and up-to-date overview of the EG control interventions worldwide.

**Methods:** We conducted a scoping review by mapping all studies on interventions for EG control following the Arksey and O’Malley Framework. We screened identified articles, and charted and coded selected papers. We classified the data based on target host, type of study, and control mechanism. We described the efficacy or safety outcomes, and the associated barriers/facilitators for the intervention. Critical appraisal was conducted.

**Results:** From 7,853 screened studies, we analyzed 45: seven centered on human interventions, 21 on animals, and 17 on both. Studies on humans focused on educational strategies and human CE monitoring. The studies on animals were field trials and most were based on Praziquantel (PZQ) for dogs. Studies focused on both animals and humans had, in general, more participants, lasted longer, and covered larger geographical areas. Overall, the quality of studies was moderate to low.

**Conclusions:** Available evidence suggests that long-term interventions aimed at both animals and humans can achieve significant reduction in EG transmission, particularly when PZQ treatment for dogs is included. Higher quality evidence, standardization of methodologies, and better reporting on post-intervention outcomes are necessary for drawing stronger conclusions. Further evidence is needed to assess the sustainability and scalability of control measures. Nonetheless, an integrative One Health approach is essential for overcoming the multiple challenges associated with sustaining long-term control efforts for Echinococcosis.

**Funding:** RCN was supported by the National Institute of Allergy and Infectious Diseases (grant nos. K01AI139284 and R01AI168291). LOC, JAB, and RCN were supported by the Fogarty International Center (grant no. D43TW012741). TAD, CG and JAB were supported by the Fogarty International Center (grant no. D43TW001140).

**Author Summary:** Cystic echinococcosis is a disease caused by the parasite *Echinococcus granulosus sensu lato*. This parasite can be found in specific areas on all continents, especially in poverty-stricken regions, increasing costs and losses. Some countries have achieved control, but most are still in the process. Our review provides a clear picture of what we currently know about these control strategies and points out where more research is needed. It highlights how the findings can improve control practices by showing what works best and address practical challenges. The review also identifies gaps in current knowledge and suggests that comparing different control methods could help find the most effective and cost-efficient solutions.

Key areas needing attention include increasing support and funding for echinococcosis, as the disease is often overlooked. More research from different fields is needed to better understand and manage the disease’s complexities. Better and ongoing surveillance is crucial for maintaining effective control strategies. Finally, comprehensive reviews that bring together findings from different studies are needed to identify what works best and improve and combine future control efforts.

## Introduction

Cystic echinococcosis (CE) is a zoonotic disease caused by the tapeworm *Echinococcus granulosus sensu lato* (EG)^1–3^. CE is most prevalent in the Mediterranean area, Eurasia, North and East Africa, and South America, where the dissemination is closely linked to home sheep slaughtering and dog feeding raw viscera^4–7^. However, human CE cases are also reported in the USA^8^, Canada^9,10^, Europe^11,12^, and Japan^13^. High infection rates among dogs, coupled with socioeconomic and ecological factors, lead to extensive human exposure^14–16^. The global burden of CE includes 184,000 disability-adjusted life years (DALYs) annually, human-associated economic losses of $764 million per year^17^, and a global cost for controlling the disease in animals and humans estimated at $3 billion^3^. Inadequate reporting of CE cases due to poor surveillance is also common globally and prevents accurate estimates of the burden of disease^18^. Despite its widespread distribution and significant global impact on human health, CE is considered a neglected tropical disease, more specifically a neglected zoonotic disease (NZD)^19–21^. CE does not receive enough attention and funding, which results in limited research efforts compared to other diseases. EG transmission involves mainly canines and ruminants, so research initiatives and intervention approaches are hindered by intersectoral work, complexities associated with ecological factors, and intricate economics related to animal husbandry^22^. Controlling NZDs offers a cost-effective chance to reduce poverty in rural regions where they are most prevalent^23^. Rural determinants of health involve social, psychological, economic and spiritual aspects that should be specifically addressed in control strategies^24^.

EG has a complex life cycle involving definitive hosts, intermediate hosts, and humans as dead-end hosts^3,25^. Intermediate hosts, primarily herbivores like sheep, develop larval cysts in organs such as the liver and lungs after ingesting pastures contaminated with parasite eggs. Definitive hosts, typically domestic or wild canids, become infected by consuming the organs of intermediate hosts containing these cysts^3,25^. In hyperendemic regions, it is common for humans to reside closely with livestock and shepherd dogs, sharing water from natural sources; however, the evidence supporting infection through contaminated water is tenuous^26–31^. In these areas, the infected shepherd dogs become the primary source of infection for both humans and livestock^6,15^.

CE control programs primarily aim to stop the transmission of the causative agent to the hosts^18,32^. Strategies targeting definite or intermediate hosts may include regular dog deworming^33,34^, dog population control^35,36^, improved management of infected viscera^37^, sheep vaccination^38^, health education programs^39^, epidemiological surveillance strategies^40–42^, improvement of diagnostic tests and infrastructure^19,43^, and integrative One Health initiatives^44^, among others. However, a common barrier to effective control programs is the lack of sufficient information needed to design, implement, and evaluate them.

A literature review is essential to identify the effective components of control and elimination interventions, aiding in strategy development and policy formulation. We conducted a comprehensive literature review, using a scoping review methodology, a useful tool in the arsenal of evidence synthesis approaches^45^. Our goal was to identify the scientific rationale, objectives, and efficacy of various interventions, providing a current overview of the current status of EG control interventions worldwide. This study provides new insights for future research and public health efforts for CE control.

## Methods

The protocol of this review is registered on the Open Science Framework (www.osf.io) with DOI 10.17605/OSF.IO/48AZR. We mapped all available evidence on interventions for EG control. We followed the Arksey and O’Malley Framework consisting of five stages: (i) Identifying the research question, (ii) Identifying relevant studies, (iii) Study selection, (iv) Charting the data, and (v) Collating, summarizing and reporting the results^46,47,45,48,49^.

### Identifying the research questions

With this review we aimed to (i) map all evidence available regarding field interventions looking to control, reduce or eliminate echinococcosis in endemic areas and, (ii) identify research gaps that can lead to future studies and, potentially, successful interventions. The following questions guided this review:

1. What strategies have been implemented, piloted, or tested to reduce, control, or eliminate EG infection in animals and/or humans?

a. What was the rationale for the study/intervention?
b. Who/what were the targets?
c. What barriers and facilitators were encountered?
2. For each intervention found in question 1

a. What was the efficacy/effectiveness outcome?
b. Was there any safety metric/outcome?
c. How long has the program/intervention worked? Is it still in place?
d. What kind of modifications/changes has the intervention undergone over time?

### Identifying relevant studies

We initially outlined a search strategy for databases and search engines for published data, followed by reference scanning and gray literature search for unpublished or difficult-to-find studies. For published data, we included MEDLINE (via PubMed), EMBASE, SCOPUS, Web of Science, Global Index Medicus and Google Scholar. The search strategy was outlined by the research team with the support of a librarian. (Appendix 1: search strategy) The search included all studies until May 2024. The results from all searches were imported and organized in Covidence™ (Melbourne, Australia), a web-based software platform that streamlines the literature review process.

### Study selection

We included all available evidence, including published, preprint, or gray literature, evaluating interventions to reduce, control, or eliminate echinococcosis by EG infections in animals and humans. Our search was global; we did not restrict studies by region. We included publications in English, Spanish, and Portuguese. Studies that included vaccines as components of field interventions were included. However, studies whose primary endpoint was to evaluate the efficacy of vaccines or deworming treatment under controlled conditions were considered out of the scope of this review and were excluded.

We screened the articles in two stages: first, by titles and abstracts, and second, by full-text. Titles and abstracts were imported into Covidence™ where reviewers assessed eligibility and filtered duplicates. The final list was evaluated by two pairs of reviewers. Both screening processes were preceded by a pilot to compare the results of the review teams, align criteria, and discuss discrepancies to reach a consensus. For quality appraisal, we used the Mixed Methods Appraisal Tool (MMAT) which allows the evaluation of multiple types of studies^50^.

### Data charting, synthesis, and reporting results

We used an electronic form/Excel® spreadsheet developed by the research team for the full-text stage for reviewers referencing, tracking, and documentation of excluded studies. Key information recorded included, but was not limited to, author(s), year of publication, methodology, and intervention characteristics. We piloted the abstraction chart to assess consistency between reviewers and calibrate the chart if needed.

The included papers were divided into types of intervention and classified according to the associated control mechanism (elimination of parasites, population control of hosts, and management of sick human/animal population, among others) and their target host. For each type of intervention, we described the efficacy or effectiveness and the safety outcome, if available, and the associated variables and the barriers/facilitators found. The results are presented in figures and tables. For studies with qualitative results, a synthesis of the main variables is presented.

### Quality appraisal

We used the Mixed Method Appraisal tool (MMAT), which allows for the appraisal stage of reviews that include qualitative, quantitative and mixed methods studies. It presents two general questions followed by five sections corresponding to different study designs, each section having five questions that evaluate quality criteria^50,51^. Each question can be answered as “yes”, “no” or “can’t tell”. The “yes” is considered as the fulfillment of the criterion. The tool does not present a cut-off point, so the score of the studies was presented descriptively as suggested by *Reporting the results of the MMAT (version 2018)*^52^ as follows:

5***** or 100% quality criteria met

4 **** or 80% quality criteria met

3 *** or 60% quality criteria met

2 ** or 40% quality criteria met

1 * or 20% quality criteria met

In the case of mixed studies, we followed the recommendation of the MMAT manual: we asked a total of 15 questions (5 questions for qualitative studies, 5 for quantitative studies and 5 for mixed studies) and considered the lowest score as the overall quality score.

## Results

We found a total of 7,853 potentially relevant studies, with 2,848 from MEDLINE, 2,139 from Scopus, 1,347 from WHO, 828 from Web of Science, 580 from Google Scholar, 108 from Embase, and 3 from gray literature. A total of 1997 duplicates were removed either manually or identified by Covidence. After screening the titles and abstracts, 309 met the eligibility criteria. Subsequently, 264 studies were excluded during full-text review, and finally, 45 studies were included in this scoping review (Fig 1). We mapped these studies across various regions globally, as depicted in Fig 3. Most of these studies were conducted in Argentina and China (n=7, each), where there are high endemicity areas. Other highly endemic areas such as Turkey and Kenya have been the subject of at least one study each. However, several other hyperendemic regions around the world have yet to be represented in the literature.

**Figure 1.**
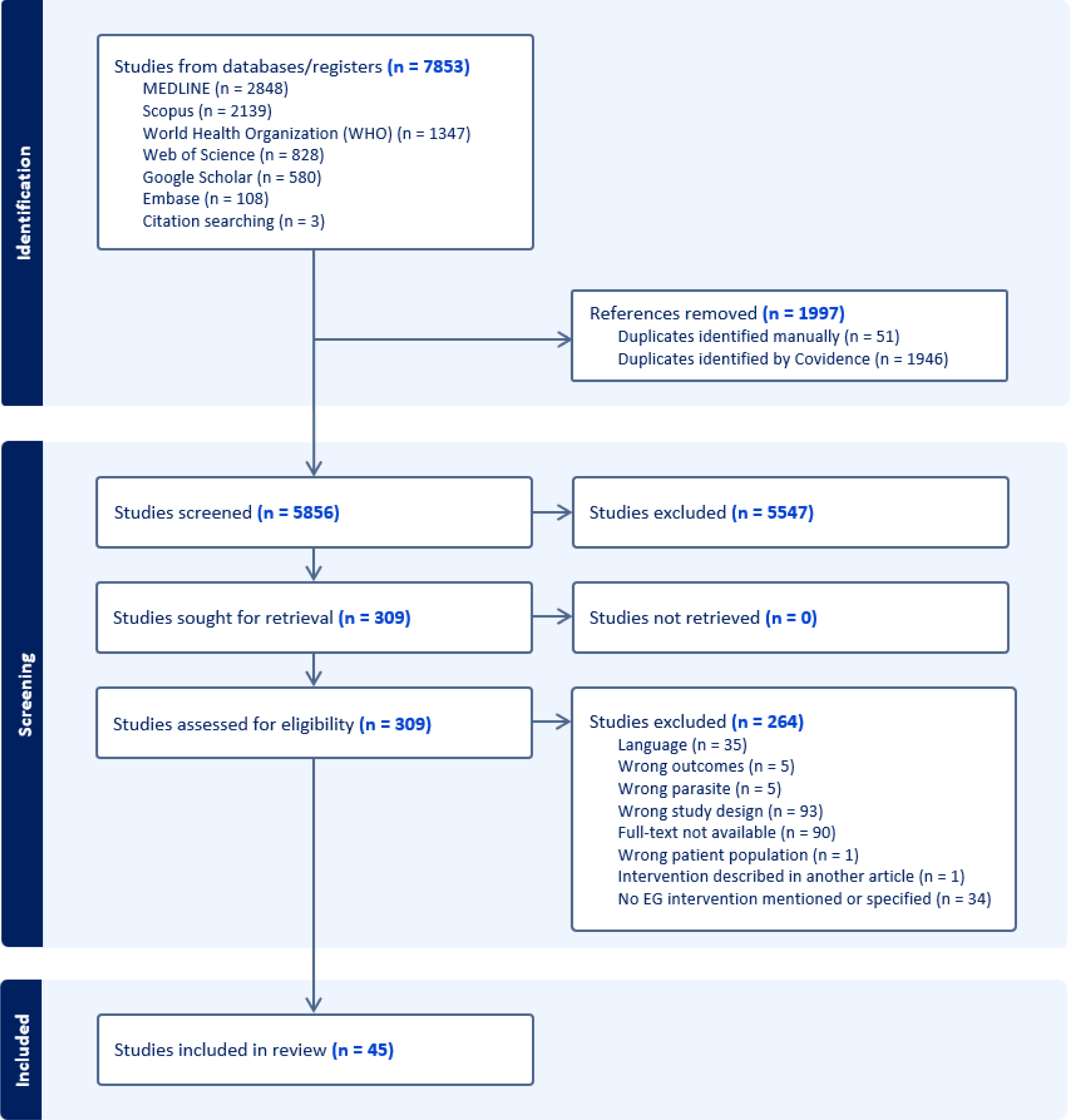
Flowchart of the inclusion/exclusion process for studies on EG control interventions.

**Figure 2.**
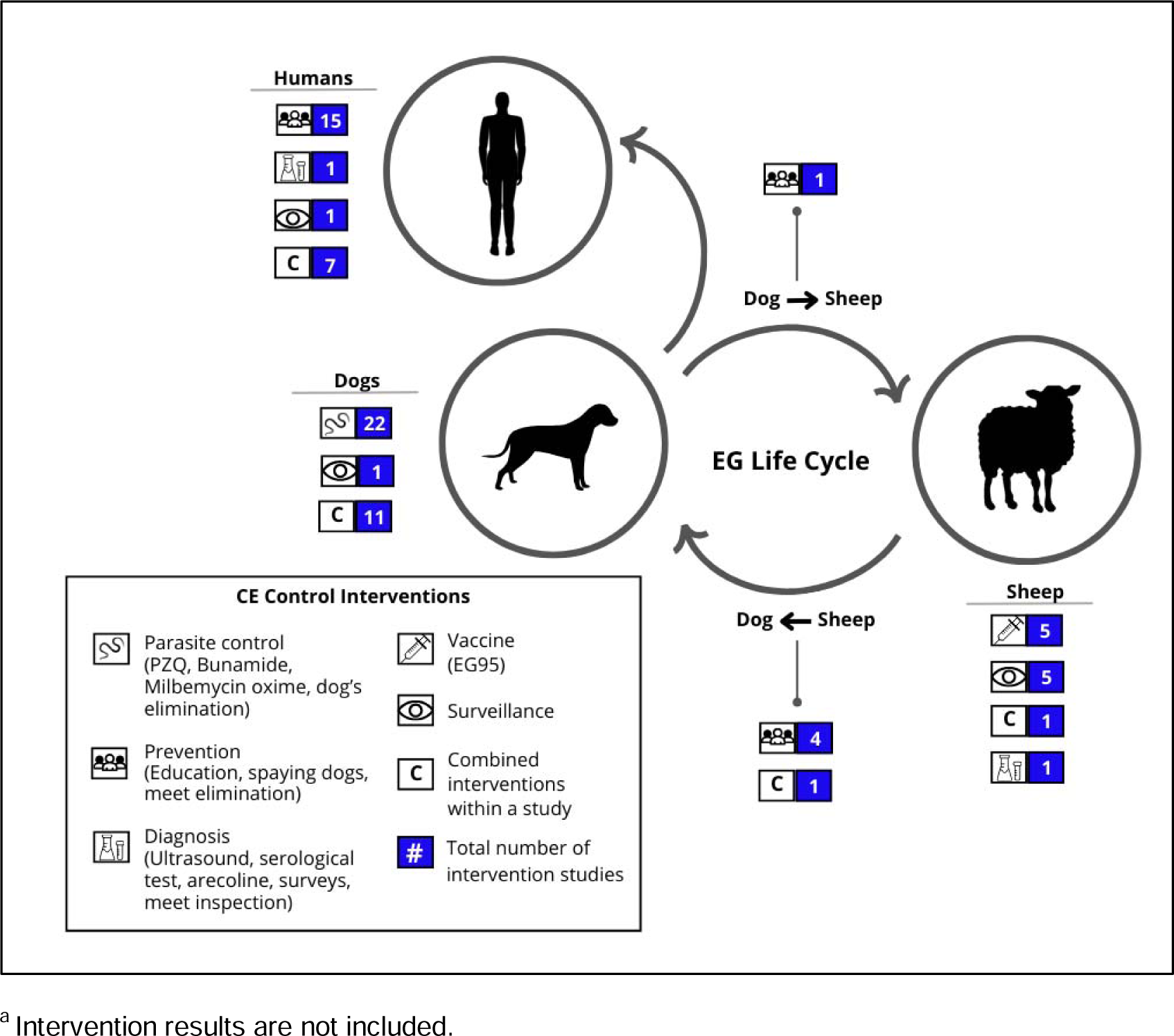
Type of control intervention reported in the selected articles and intervention target within the EG life cycle.^a^.

**Figure 3.**
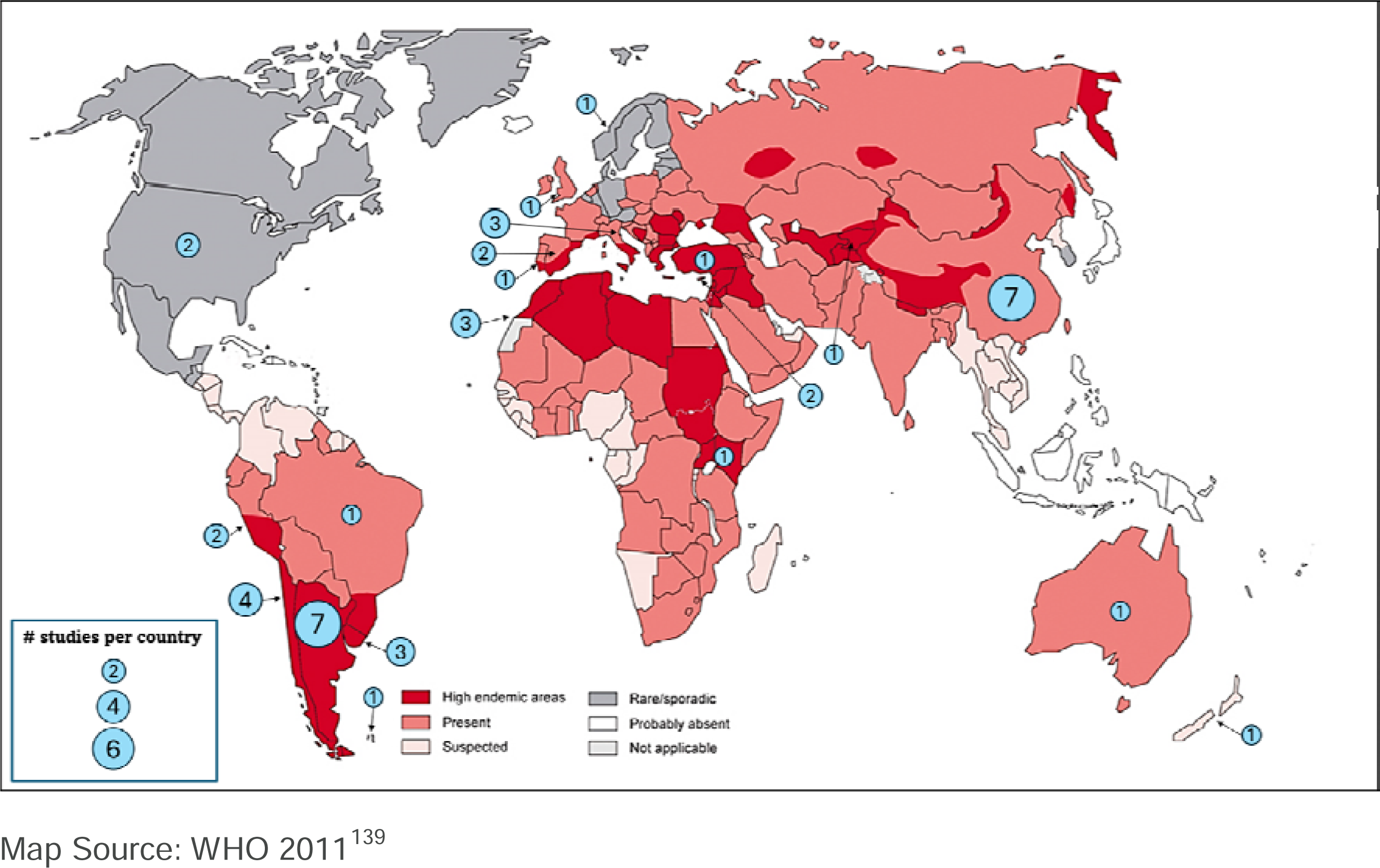
Global distribution of EG and cystic echinococcosis and studies.

### Interventions in humans

Of the 45 articles reviewed, seven studies reported interventions aimed only at humans (Table 1a). Six studies primarily focused on education and awareness and one study focused on CE monitoring. In Portugal, a 13-year program unveiled 62 CE cases via ultrasound, 24 cases via serology, and 21 cases via both. Health information was provided to adults, with specific activities designed for children. CE prevalence decreased by 20% midway through the study and by the end, feeding raw viscera to dogs dropped from 60% to 20%^53^. In Peru, nine educational sessions were applied to 28 school children. A high-level knowledge of CE increased from 50% to 100% in the intervention group^54^. In Chile, a 7-level educational intervention run over 4 years for 3,145 participants aged 2 to 20 improved knowledge at all levels. Compared to urban, rural schools showed greater improvement. The biggest changes were in knowledge about hand washing, washing fruits and vegetables, and avoiding dog licking. Knowledge of responsible dog ownership was highest before the intervention and showed the least change^55^. In the USA, a 73-item questionnaire was applied to evaluate an educational program, including knowledge and the willingness to engage in preventive actions. In 8 years, awareness increased from 28% to 83%^56^. In Turkey, awareness interventions were applied to children, the general public, and health care professionals. In children, knowledge increased from 5.8% to almost 90%. No efficacy measure is provided for adults^39^. In Morocco, an integrated health messaging intervention was implemented including EG, rabies, and other zoonoses. “Piggy-backing” on high priority diseases like rabies allowed participants to better retain information on EG^57^. In Argentina, an ultrasound monitoring trial in humans was implemented to support a comprehensive control program. The trial revealed that prevalence reduced from 5.5% to 4.04%. Additionally, high-risk groups were identified and this evidence informed program actions^58^. Details about these interventions focused on humans are found in Table 1a.

**Table 1a.** Interventions in Humans.

| # | Author, year | Country of study | Study design | Sample size | Inclusion criteria | Main objective | Intervention | Intervention duration | Main outcomes |
| --- | --- | --- | --- | --- | --- | --- | --- | --- | --- |
| **Trials** |  |  |  |  |  |  |  |  |  |
| 1 | **Larrieu E, et al., 1989** | Argentina | Field trial | 689 people. | 542 asymptomatic volunteers and 147 asymptomatic cohabitants of CE cases. | Control and prevention | Portable ultrasound screening in general public and cohabitants of CE cases that overwent surgery, with a 3-year gap during the national control program. Prevalence was reported. | 3 years (1984-1986) | Prevalence of CE. |
| 2 | **Hernández Mira GM, et al., 2008** | Portugal | Field trial | 2,152 people. | Rural volunteers: 780 men and 1362 women. | Eradicate | Tracking of CE cases in three phases.<br>- Phase 1: Epidemiological questionnaires to volunteers.<br>- Phase 2: Obtaining a blood samples for serological testing.<br>- Phase 3: Abdominal ultrasound. This actions included the distribution of informative brochures along with explanation of their content, small informational sessions for general population and specific activities for school-age children. | 13 years (1994-2006) | Prevalence of CE. Change in people habits. |
| 3 | **Pariona-Díaz A, et al., 2018** | Peru | Pre post | 56 people. | Experimental and control group, both of 28 boys and girls from 5th grade from a public school in Huancavelica. | Prevention | Educative program: 9 in-person sessions of 2.5 hours weekly. The program, using the Bustos operative didactic model, employed various techniques and teaching materials like small group sizes, guidance, active student participation throughout the learning sessions, self-correction of answers by students, and reinforcement at the end of each session. After the intervention a questionnaire of 40 items was applied to assess effectivity. | 9-10 weeks | Knowledge level about CE after the implementation of the educational program. |
| 4 | **Castillo-Montes M, et al., 2023** | Chile | Pre post | 3145 people. | Students from preschool, primary and secondary. All schools from three communes were included. | Prevention and Control | Educative program: First, areas of knowledge about CE were defined and seven educational levels were differentiated by grade of education. A didactic guide with different objectives and methodologies was assigned to each level. Teachers were trained to develop the program in three sessions (initial, profundization, closure). Seven pre and post tests previously validated were applied before the first and after the last session. | 5 years (2014-2018) | Changes in the knowledge level about CE after the educational program. |
| **Observational** |  |  |  |  |  |  |  |  |  |
| 5 | **Condie S, et al., 1981** | USA | Cross-sectional | People from 156 households. | Adult members of each family from Fountain Green, Utah. | Prevention and control | A 73-item questionnaire was constructed to investigate the impact of an educational-control program. The questionnaire included information about CE and the relationship between antecedent variables and the willingness to engage in six preventive actions: medicating dogs regularly, attending screening clinics, tying up their dogs, preventing dogs from eating sheep liver and lungs, disposing sheep carcasses in covered pits and bury or burn sheep carcasses. | 3 weeks (August 1978) | Correlation of attitudes and knowledge, and the willingness to participate in the public health control program. |
| 6 | **Ducrotoy MJ, et al., 2015** | Morocco | Convergent | 656 people for focus groups.<br>6910 people for evaluation questionnaire and pop quiz.<br>33 teachers for evaluation forms.<br>23 teachers for interviews. | Three groups in rural areas of Sidi Kacem province:<br>- Community, including women, men and children,<br>- School children, and,<br>- Stakeholders, including NGOs, healthcare professionals and local governments. | Prevention | For the community group, educational events focused on rabies, CE, leishmaniasis, brucellosis and bovine tuberculosis. They tried different combinations of these diseases. For events that focus solely on bacterial zoonosis, there was a separation between adults of 21 communes and school children from 23 schooled-bases. Education of stakeholders took place in participatory workshops and 4 education sessions of integrated messaging on all five zoonosis. Evaluation was done in focus groups, post presentation pop quizzes, and evaluation questionnaires. In schools, evaluation forms were given to teachers and headmasters, and informal interviews were held with teachers. | 18 months | Pros and cons of the integrated health messaging intervention. |
| 7 | Altintas NA, et al., 2021 | Turkey | Descriptive | 5208 people. | Three target groups from CE endemic districts:<br>- Elementary school 3rd and 4th year students, teachers and administrators from schools,<br>- General public, and,<br>- Healthcare professionals. | Prevention | Awareness activities:<br>Student adapted presentations were given in 26 schools for students, teachers and administrators. Brochures and posters were distributed.<br>For general public seminars and brochure distribution about CE. Training of healthcare professionals included presentations and distribution of brochures. | 1 year (2019-2020) | Number of attendees to the seminars.<br>Knowledge change in school children. |

\*CE: cystic echinococcosis, EG: echinococcus granulosus

### Interventions in animals

We reviewed 21 articles of interventions aimed only at animals (Table 1b). Six interventions targeted both dogs and sheep, 12 targeted only dogs, and 3 only sheep. Most studies were based on the delivery of Praziquantel (PZQ). In Australia, an intervention included testing dogs, treating positive dogs with bunamide at days 0 and 14, and recommendations for home slaughtering. The incidence in dogs dropped from 11.3% to 2.1% in a 4-year period^59^. In Uruguay, different PZQ frequencies in dogs were compared. One year of PZQ treatment every 6 weeks reduced the prevalence in lambs to 0, while in the 12- and 16-week schemes, as well as in the control group, the prevalence ranged from 4.3 to 18.6%^60^. Also in Uruguay, a PZQ-based 9-years control program in dogs reduced the prevalence in dogs from 22.7% to 1.5% and, in 4-year-old sheep from 49.3% to 18.5%^61^. Similarly in Brazil and China, dogs that were treated with PZQ every 30 days had their prevalence reduced from 10.67% to 0.74%^62^ and from 7.3% to 1.7%^63^ respectively. In Peru, an interesting PZQ regimen was implemented: two treatment cycles were administered to dogs with a 12-month interval between them. In each cycle, dogs received three doses of PZQ, one dose per month. Prevalence in dogs reduced from 47.2% to 1.3%. Unfortunately, prevalence was only evaluated once one month after the last dose^64^. In Argentina, a control program that started in 1980 had multiple modifications. It began with an EG baseline for dogs, sheep, and the environment. Later lambs received EG95 vaccine and multiple boosters. Female sheep got an extra dose for colostral immunity. After 3 years, prevalence in sheep dropped from 26.2% to 7.8%^65^. This program, updated in 2009, was later evaluated through copro ELISA in dogs, necropsy of adult sheep, and ultrasound screening in children. By 2017, infection in 6 y.o. sheep decreased from 56.3% to 21.6%, in dogs from 9.6% to 3.7%, and in children from 38 cases to only one^66^. In the same program, between 2018-2022, in addition to EG95 in sheep, dogs were treated with PZQ 4-times a year. Prevalence among vaccinated and unvaccinated sheep was 21% and 66%, respectively^67^. In Chile, a neighboring country to Argentina, a 3-year program vaccinated sheep with EG95 vaccine at 2 and 3 months old, and then annually. By the end of the program, cyst fertility dropped from 28.1% to 8%^68^. Another multi-pronged intervention was conducted in Morocco, a 4-year program compared PZQ for dogs every 4 months, EG95 vaccination of lambs, and both combined. Dog prevalence dropped from 35% to 9%. Post-intervention, the prevalence of viable cysts in sheep was 5% with combined intervention, 8% with sheep vaccination, 69% with dog treatment only, and 77% with no intervention^69^.

**Table 1b.**
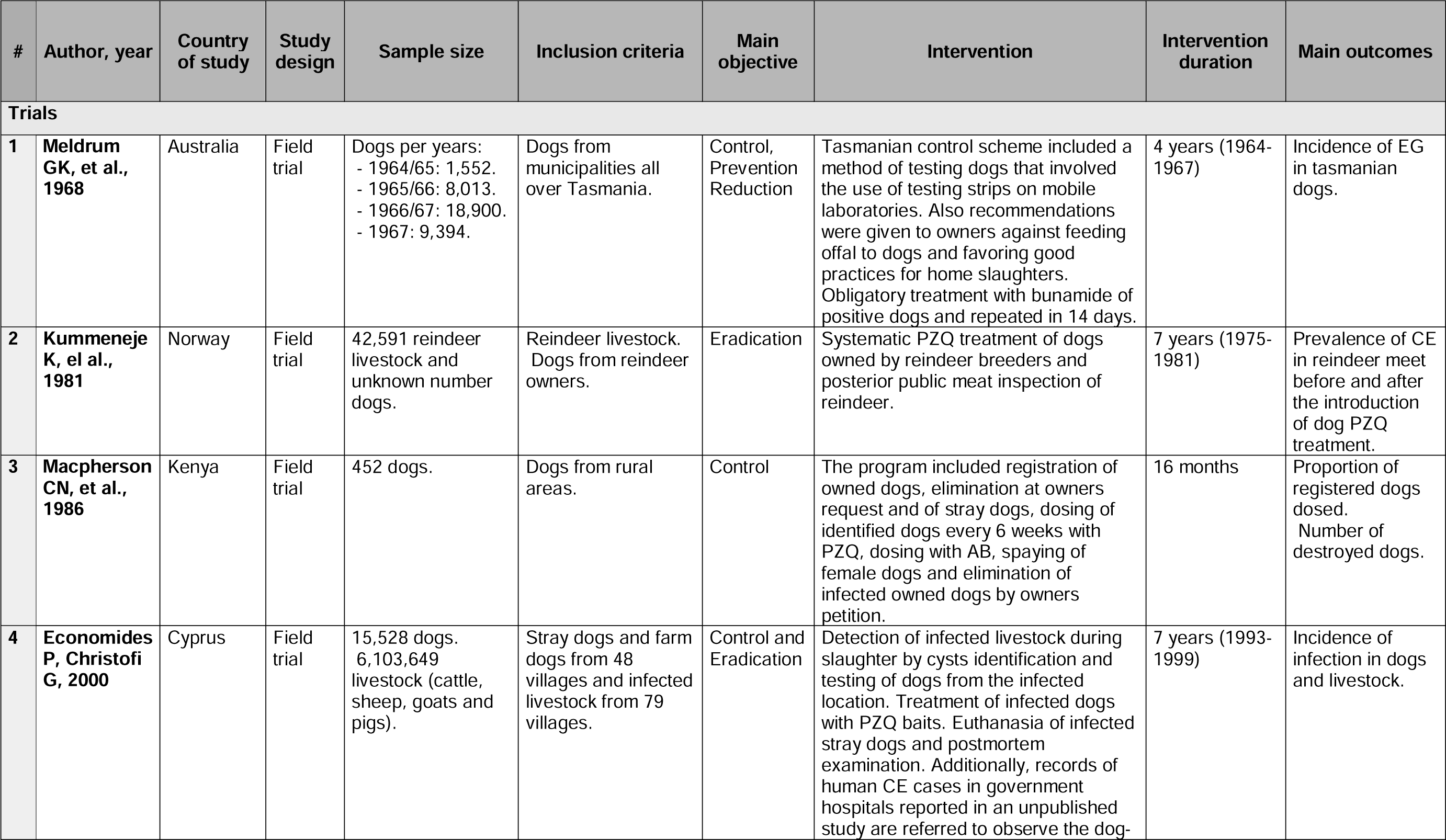
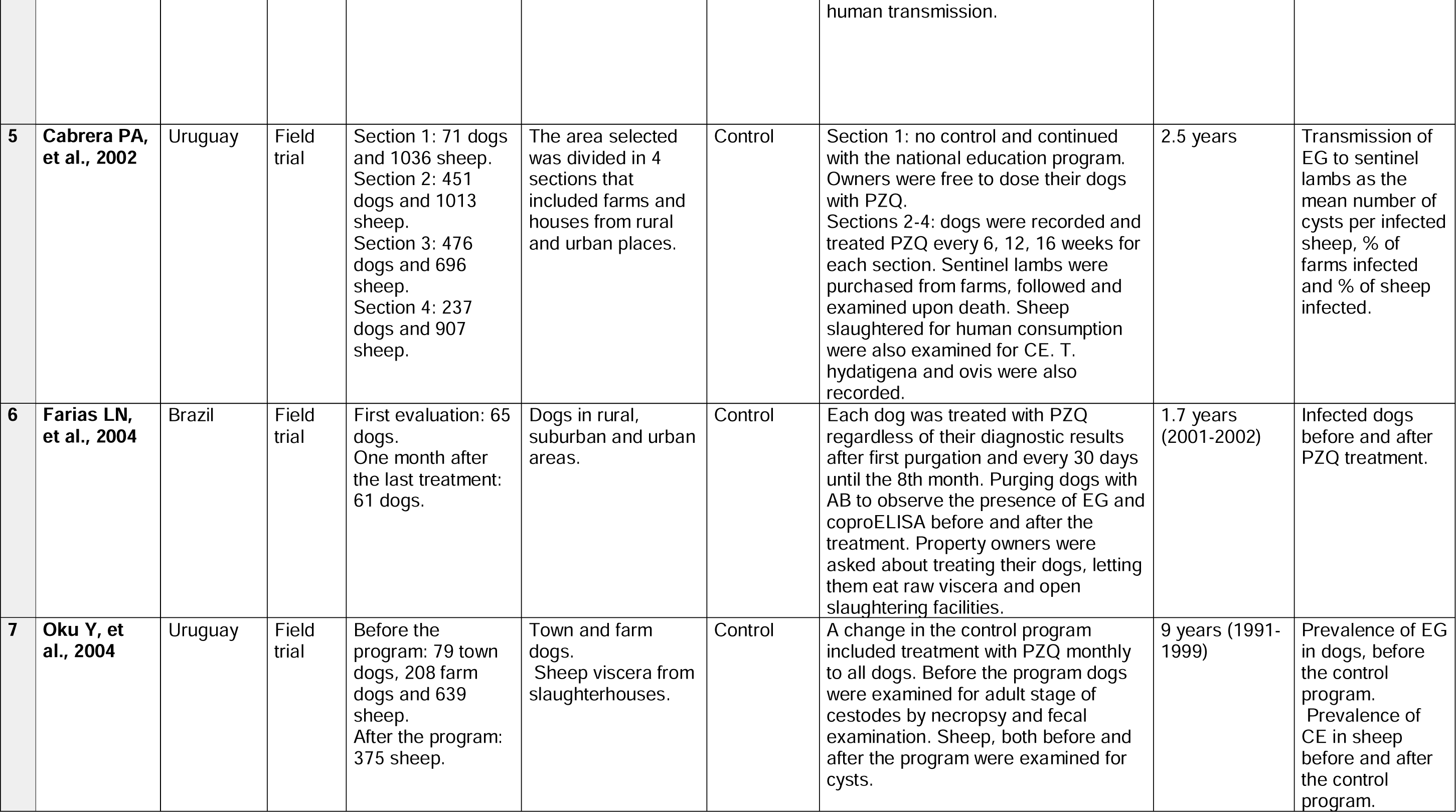
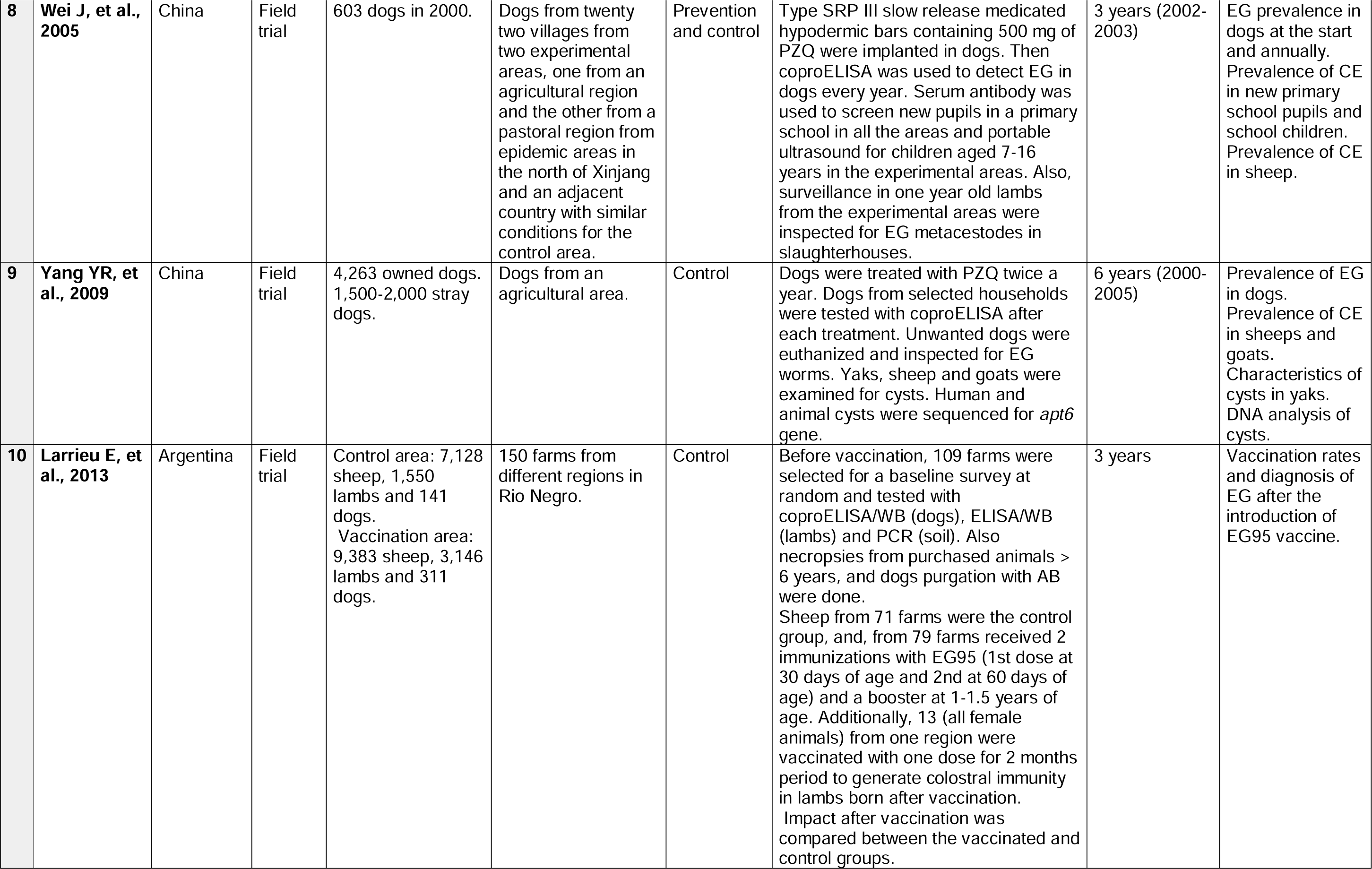
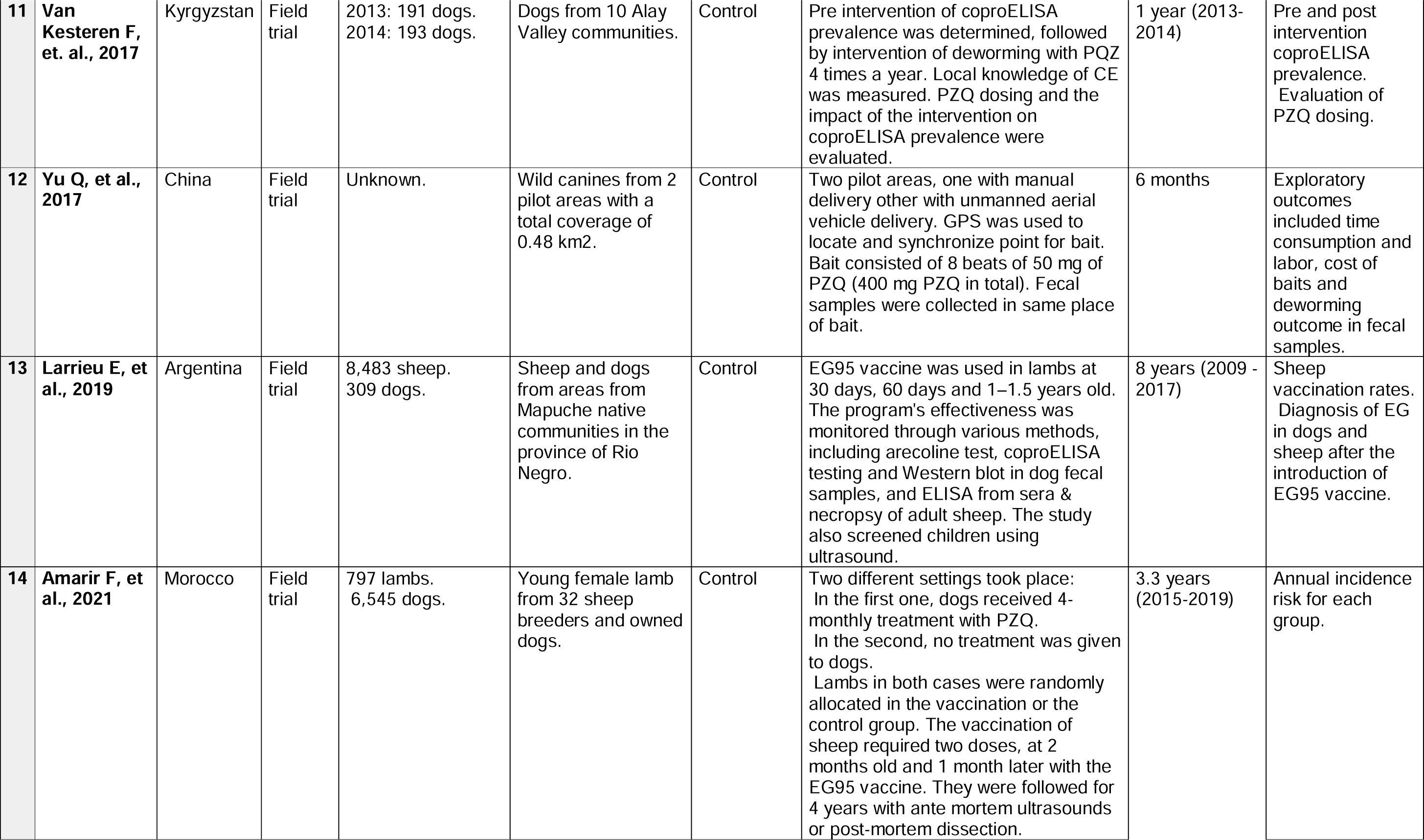
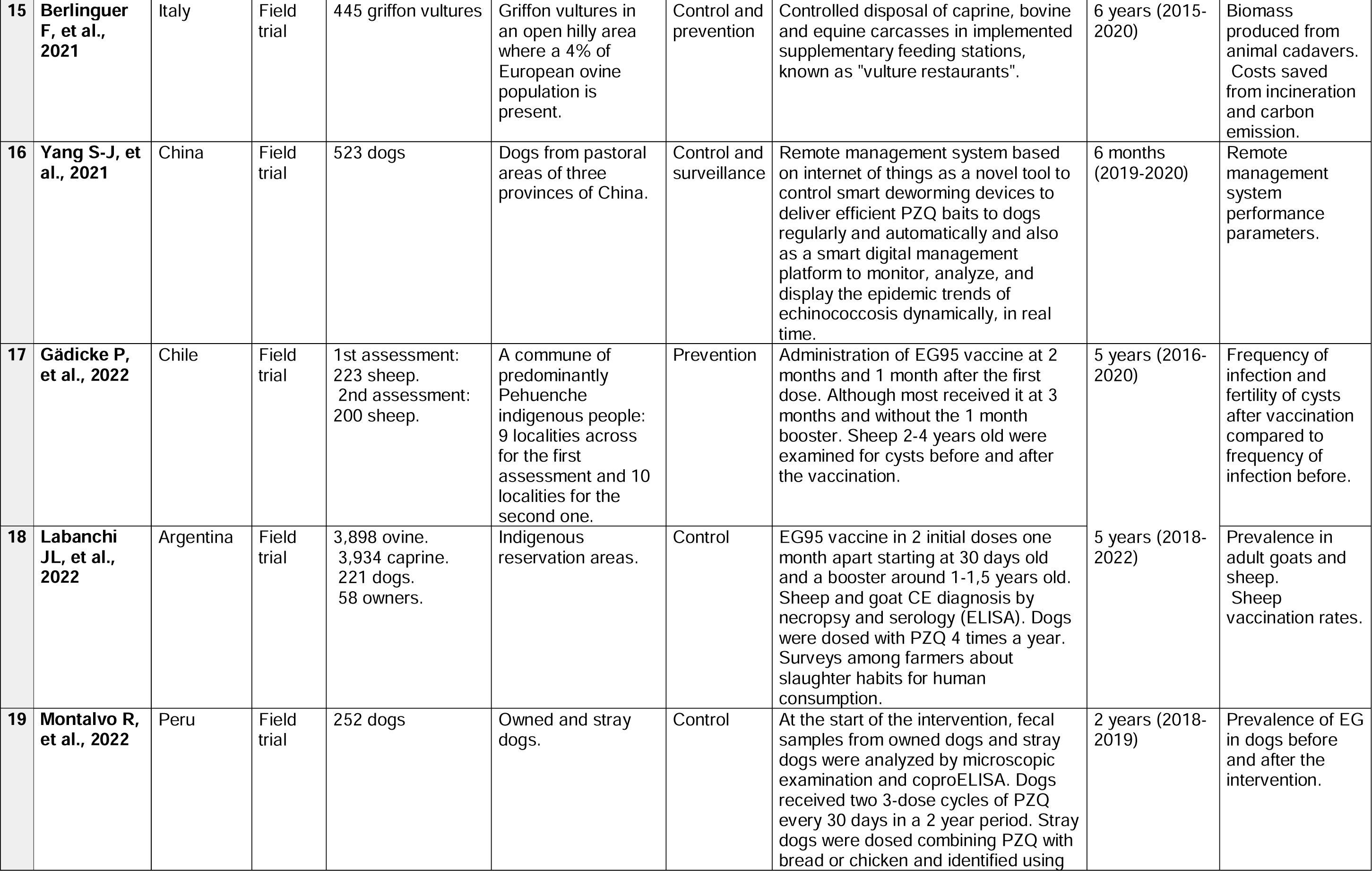
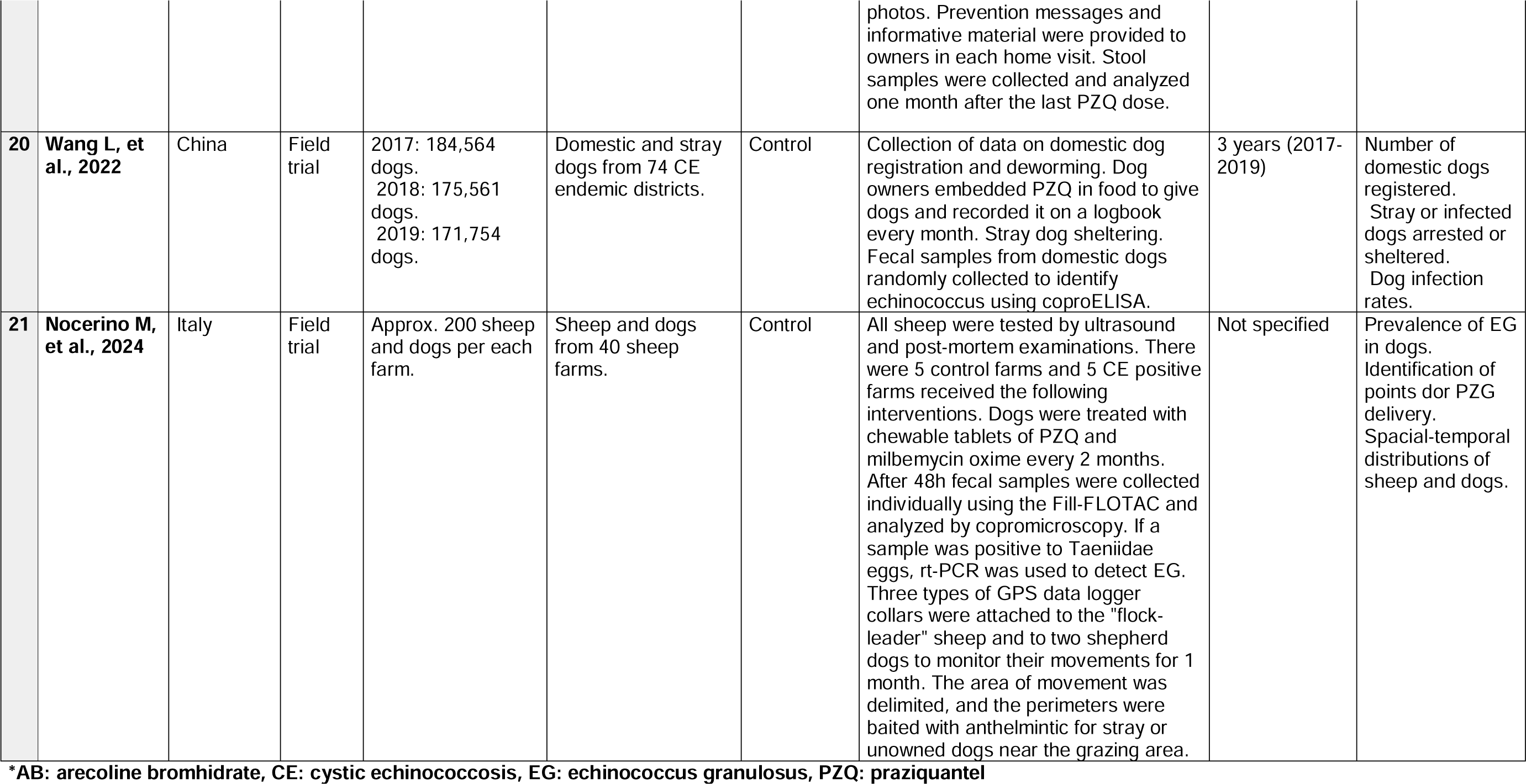
Interventions in Animals.

| # | Author, year | Country of study | Study design | Sample size | Inclusion criteria | Main objective | Intervention | Intervention duration | Main outcomes |
| --- | --- | --- | --- | --- | --- | --- | --- | --- | --- |
| **Trials** |  |  |  |  |  |  |  |  |  |
| 1 | **Meldrum GK, et al., 1968** | Australia | Field trial | Dogs per years:<br>- 1964/65: 1,552.<br>- 1965/66: 8,013.<br>- 1966/67: 18,900.<br>- 1967: 9,394. | Dogs from municipalities all over Tasmania. | Control, Prevention Reduction | Tasmanian control scheme included a method of testing dogs that involved the use of testing strips on mobile laboratories. Also recommendations were given to owners against feeding offal to dogs and favoring good practices for home slaughters. Obligatory treatment with bunamide of positive dogs and repeated in 14 days. | 4 years (1964-1967) | Incidence of EG in tasmanian dogs. |
| 2 | **Kummeneje K, et al., 1981** | Norway | Field trial | 42,591 reindeer livestock and unknown number dogs. | Reindeer livestock. Dogs from reindeer owners. | Eradication | Systematic PZQ treatment of dogs owned by reindeer breeders and posterior public meat inspection of reindeer. | 7 years (1975-1981) | Prevalence of CE in reindeer meet before and after the introduction of dog PZQ treatment. |
| 3 | **Macpherson CN, et al., 1986** | Kenya | Field trial | 452 dogs. | Dogs from rural areas. | Control | The program included registration of owned dogs, elimination at owners request and of stray dogs, dosing of identified dogs every 6 weeks with PZQ, dosing with AB, spaying of female dogs and elimination of infected owned dogs by owners petition. | 16 months | Proportion of registered dogs dosed. Number of destroyed dogs. |
| 4 | **Economides P, Christofi G, 2000** | Cyprus | Field trial | 15,528 dogs. 6,103,649 livestock (cattle, sheep, goats and pigs). | Stray dogs and farm dogs from 48 villages and infected livestock from 79 villages. | Control and Eradication | Detection of infected livestock during slaughter by cysts identification and testing of dogs from the infected location. Treatment of infected dogs with PZQ baits. Euthanasia of infected stray dogs and postmortem examination. Additionally, records of human CE cases in government hospitals reported in an unpublished study are referred to observe the dog- human transmission. | 7 years (1993-1999) | Incidence of infection in dogs and livestock. |
| 5 | **Cabrera PA, et al., 2002** | Uruguay | Field trial | Section 1: 71 dogs and 1036 sheep.<br>Section 2: 451 dogs and 1013 sheep.<br>Section 3: 476 dogs and 696 sheep.<br>Section 4: 237 dogs and 907 sheep. | The area selected was divided in 4 sections that included farms and houses from rural and urban places. | Control | Section 1: no control and continued with the national education program. Owners were free to dose their dogs with PZQ.<br>Sections 2-4: dogs were recorded and treated PZQ every 6, 12, 16 weeks for each section. Sentinel lambs were purchased from farms, followed and examined upon death. Sheep slaughtered for human consumption were also examined for CE. T. hydatigena and ovis were also recorded. | 2.5 years | Transmission of EG to sentinel lambs as the mean number of cysts per infected sheep, % of farms infected and % of sheep infected. |
| 6 | **Farias LN, et al., 2004** | Brazil | Field trial | First evaluation: 65 dogs.<br>One month after the last treatment: 61 dogs. | Dogs in rural, suburban and urban areas. | Control | Each dog was treated with PZQ regardless of their diagnostic results after first purgation and every 30 days until the 8th month. Purging dogs with AB to observe the presence of EG and coproELISA before and after the treatment. Property owners were asked about treating their dogs, letting them eat raw viscera and open slaughtering facilities. | 1.7 years (2001-2002) | Infected dogs before and after PZQ treatment. |
| 7 | **Oku Y, et al., 2004** | Uruguay | Field trial | Before the program: 79 town dogs, 208 farm dogs and 639 sheep.<br>After the program: 375 sheep. | Town and farm dogs.<br>Sheep viscera from slaughterhouses. | Control | A change in the control program included treatment with PZQ monthly to all dogs. Before the program dogs were examined for adult stage of cestodes by necropsy and fecal examination. Sheep, both before and after the program were examined for cysts. | 9 years (1991-1999) | Prevalence of EG in dogs, before the control program.<br>Prevalence of CE in sheep before and after the control program. |
| 8 | Wei J, et al., 2005 | China | Field trial | 603 dogs in 2000. | Dogs from twenty two villages from two experimental areas, one from an agricultural region and the other from a pastoral region from epidemic areas in the north of Xinjiang and an adjacent country with similar conditions for the control area. | Prevention and control | Type SRP III slow release medicated hypodermic bars containing 500 mg of PZQ were implanted in dogs. Then coproELISA was used to detect EG in dogs every year. Serum antibody was used to screen new pupils in a primary school in all the areas and portable ultrasound for children aged 7-16 years in the experimental areas. Also, surveillance in one year old lambs from the experimental areas were inspected for EG metacestodes in slaughterhouses. | 3 years (2002-2003) | EG prevalence in dogs at the start and annually. Prevalence of CE in new primary school pupils and school children. Prevalence of CE in sheep. |
| 9 | Yang YR, et al., 2009 | China | Field trial | 4,263 owned dogs. 1,500-2,000 stray dogs. | Dogs from an agricultural area. | Control | Dogs were treated with PZQ twice a year. Dogs from selected households were tested with coproELISA after each treatment. Unwanted dogs were euthanized and inspected for EG worms. Yaks, sheep and goats were examined for cysts. Human and animal cysts were sequenced for *apt6* gene. | 6 years (2000-2005) | Prevalence of EG in dogs. Prevalence of CE in sheeps and goats. Characteristics of cysts in yaks. DNA analysis of cysts. |
| 10 | Larrieu E, et al., 2013 | Argentina | Field trial | Control area: 7,128 sheep, 1,550 lambs and 141 dogs.<br>Vaccination area: 9,383 sheep, 3,146 lambs and 311 dogs. | 150 farms from different regions in Rio Negro. | Control | Before vaccination, 109 farms were selected for a baseline survey at random and tested with coproELISA/WB (dogs), ELISA/WB (lambs) and PCR (soil). Also necropsies from purchased animals > 6 years, and dogs purgation with AB were done.<br>Sheep from 71 farms were the control group, and, from 79 farms received 2 immunizations with EG95 (1st dose at 30 days of age and 2nd at 60 days of age) and a booster at 1-1.5 years of age. Additionally, 13 (all female animals) from one region were vaccinated with one dose for 2 months period to generate colostral immunity in lambs born after vaccination.<br>Impact after vaccination was compared between the vaccinated and control groups. | 3 years | Vaccination rates and diagnosis of EG after the introduction of EG95 vaccine. |
| 11 | **Van Kesteren F, et. al., 2017** | Kyrgyzstan | Field trial | 2013: 191 dogs.<br>2014: 193 dogs. | Dogs from 10 Alay Valley communities. | Control | Pre intervention of coproELISA prevalence was determined, followed by intervention of deworming with PQZ 4 times a year. Local knowledge of CE was measured. PZQ dosing and the impact of the intervention on coproELISA prevalence were evaluated. | 1 year (2013-2014) | Pre and post intervention coproELISA prevalence. Evaluation of PZQ dosing. |
| 12 | **Yu Q, et al., 2017** | China | Field trial | Unknown. | Wild canines from 2 pilot areas with a total coverage of 0.48 km<sup>2</sup>. | Control | Two pilot areas, one with manual delivery other with unmanned aerial vehicle delivery. GPS was used to locate and synchronize point for bait. Bait consisted of 8 beats of 50 mg of PZQ (400 mg PZQ in total). Fecal samples were collected in same place of bait. | 6 months | Exploratory outcomes included time consumption and labor, cost of baits and deworming outcome in fecal samples. |
| 13 | **Larrieu E, et al., 2019** | Argentina | Field trial | 8,483 sheep.<br>309 dogs. | Sheep and dogs from areas from Mapuche native communities in the province of Rio Negro. | Control | EG95 vaccine was used in lambs at 30 days, 60 days and 1–1.5 years old. The program's effectiveness was monitored through various methods, including arecoline test, coproELISA testing and Western blot in dog fecal samples, and ELISA from sera & necropsy of adult sheep. The study also screened children using ultrasound. | 8 years (2009 - 2017) | Sheep vaccination rates. Diagnosis of EG in dogs and sheep after the introduction of EG95 vaccine. |
| 14 | **Amarir F, et al., 2021** | Morocco | Field trial | 797 lambs.<br>6,545 dogs. | Young female lamb from 32 sheep breeders and owned dogs. | Control | Two different settings took place:<br>In the first one, dogs received 4-monthly treatment with PZQ.<br>In the second, no treatment was given to dogs.<br>Lambs in both cases were randomly allocated in the vaccination or the control group. The vaccination of sheep required two doses, at 2 months old and 1 month later with the EG95 vaccine. They were followed for 4 years with ante mortem ultrasounds or post-mortem dissection. | 3.3 years (2015-2019) | Annual incidence risk for each group. |
| 15 | **Berlinguer F, et al., 2021** | Italy | Field trial | 445 griffon vultures | Griffon vultures in an open hilly area where a 4% of European ovine population is present. | Control and prevention | Controlled disposal of caprine, bovine and equine carcasses in implemented supplementary feeding stations, known as "vulture restaurants". | 6 years (2015-2020) | Biomass produced from animal cadavers. Costs saved from incineration and carbon emission. |
| 16 | **Yang S-J, et al., 2021** | China | Field trial | 523 dogs | Dogs from pastoral areas of three provinces of China. | Control and surveillance | Remote management system based on internet of things as a novel tool to control smart deworming devices to deliver efficient PZQ baits to dogs regularly and automatically and also as a smart digital management platform to monitor, analyze, and display the epidemic trends of echinococcosis dynamically, in real time. | 6 months (2019-2020) | Remote management system performance parameters. |
| 17 | **Gädicke P, et al., 2022** | Chile | Field trial | 1st assessment: 223 sheep. 2nd assessment: 200 sheep. | A commune of predominantly Pehuenche indigenous people: 9 localities across for the first assessment and 10 localities for the second one. | Prevention | Administration of EG95 vaccine at 2 months and 1 month after the first dose. Although most received it at 3 months and without the 1 month booster. Sheep 2-4 years old were examined for cysts before and after the vaccination. | 5 years (2016-2020) | Frequency of infection and fertility of cysts after vaccination compared to frequency of infection before. |
| 18 | **Labanchi JL, et al., 2022** | Argentina | Field trial | 3,898 ovine. 3,934 caprine. 221 dogs. 58 owners. | Indigenous reservation areas. | Control | EG95 vaccine in 2 initial doses one month apart starting at 30 days old and a booster around 1-1,5 years old. Sheep and goat CE diagnosis by necropsy and serology (ELISA). Dogs were dosed with PZQ 4 times a year. Surveys among farmers about slaughter habits for human consumption. | 5 years (2018-2022) | Prevalence in adult goats and sheep. Sheep vaccination rates. |
| 19 | **Montalvo R, et al., 2022** | Peru | Field trial | 252 dogs | Owned and stray dogs. | Control | At the start of the intervention, fecal samples from owned dogs and stray dogs were analyzed by microscopic examination and coproELISA. Dogs received two 3-dose cycles of PZQ every 30 days in a 2 year period. Stray dogs were dosed combining PZQ with bread or chicken and identified using photos. Prevention messages and informative material were provided to owners in each home visit. Stool samples were collected and analyzed one month after the last PZQ dose. | 2 years (2018-2019) | Prevalence of EG in dogs before and after the intervention. |
| 20 | Wang L, et al., 2022 | China | Field trial | 2017: 184,564 dogs.<br>2018: 175,561 dogs.<br>2019: 171,754 dogs. | Domestic and stray dogs from 74 CE endemic districts. | Control | Collection of data on domestic dog registration and deworming. Dog owners embedded PZQ in food to give dogs and recorded it on a logbook every month. Stray dog sheltering. Fecal samples from domestic dogs randomly collected to identify echinococcus using coproELISA. | 3 years (2017-2019) | Number of domestic dogs registered.<br>Stray or infected dogs arrested or sheltered.<br>Dog infection rates. |
| 21 | Nocerino M, et al., 2024 | Italy | Field trial | Approx. 200 sheep and dogs per each farm. | Sheep and dogs from 40 sheep farms. | Control | All sheep were tested by ultrasound and post-mortem examinations. There were 5 control farms and 5 CE positive farms received the following interventions. Dogs were treated with chewable tablets of PZQ and milbemycin oxime every 2 months. After 48h fecal samples were collected individually using the Fill-FLOTAC and analyzed by copromicroscopy. If a sample was positive to Taeniidae eggs, rt-PCR was used to detect EG. Three types of GPS data logger collars were attached to the "flock-leader" sheep and to two shepherd dogs to monitor their movements for 1 month. The area of movement was delimited, and the perimeters were baited with anthelmintic for stray or unowned dogs near the grazing area. | Not specified | Prevalence of EG in dogs.<br>Identification of points for PZQ delivery.<br>Spatial-temporal distributions of sheep and dogs. |

\*AB: arecoline bromhidrate, CE: cystic echinococcosis, EG: echinococcus granulosus, PZQ: praziquantel

Cyprus and Norway provided data from hypoendemic areas. In Norway, dogs were treated with PZQ biannually first and annually in the following years. After 4 years, the prevalence in reindeer reduced from 1.5% to 0.11%^70^. In Cyprus, infected livestock were identified in slaughterhouses, dogs were tested using arecoline and coproELISA, and infected dogs were euthanized and examined or treated with PZQ. Sheep prevalence reduced from 0.033% to 0.007%^71^. Several studies did not report efficacy metrics or reported negative results. In Kenya, a 16-month control program included dog population control, administering arecoline, and treating infected dogs with PZQ every six weeks. Prevalence reduced but remained high and returned to pre-control levels within three months^72^. In Kyrgyzstan, dogs were treated with PZQ four times for a year. Pre-intervention coproELISA prevalence was 20.1%; however, post-intervention prevalence was not reported^73^. In China, multiple studies have been reported. Baits with PZQ were tested to treat dogs and other canids every other month; however, after 7 months, no significant effect was detected in high-prevalence areas^74^. In Buddhist areas, where dog culling is not accepted, administering PZQ twice a year for five years reduced EG prevalence in dogs only from 27% to 20%^75^. In a different study, after 3 years of implanting slow-released PZQ-medicated bars in dogs, copro ELISA positivity in dogs plummeted from 41.2% to 3%, seropositivity in children reduced from 41.2% to 5.4%, and prevalence in sheep dropped from 44.8% to 10.7%^76^. Some interventions have explored innovative approaches. In China, smart collars enabled automatic delivery of PZQ baits to dogs. Collars also had sensors to provide real-time data that could be used to study trends of echinococcosis. Post-intervention prevalence was not provided^77^. In Italy, carcass disposal was enhanced through vulture feeding stations. CE prevalence in sheep was 65.3% and, though no post-intervention estimates are provided, the authors concluded that the program reduced CE risk and burden^78^. Also in Italy, GPS data loggers were attached to a sheep leader and two shepherd dogs to delimit grazing areas. Grazing areas’ perimeters were baited with anthelmintics for free-roaming dogs, proposing another method of control^79^. Details of these interventions focused on animals can be found in Table 1b.

### Interventions in animals and humans

We reviewed 17 articles focused on both animals and humans (Table 1c). Multiple long-term programs and short-term studies are reported from South America and described integrated interventions dosing dogs, vaccinating sheep, educating and screening humans, and enhancing abattoir surveillance. In Argentina, dogs received PZQ more frequently in high-risk areas and were monitored using arecoline and in-situ analysis. Education was provided to schools, homes, and to rural populations through accessible media. Sheep were inspected at abattoirs and surveillance data collection improved. After two years, dog prevalence decreased from 31.5% to 4.24%^80^. After 17 years of implementation, new human cases in children under 11 decreased by 77%^81^. Also in Argentina, ultrasound surveys in children and copro ELISA surveys in dogs were used to identify hotspots of transmission and strengthen control program activities (education, diagnosis and treatment in humans, deworming of dogs, and vaccination of sheep). Dog prevalence reduced from 32% to 15.6%^82^. In Uruguay, a rural intervention included monthly PZQ plus broad-spectrum anthelmintics 1 to 3 times a year, copro ELISA diagnosis, free dog castration, and ultrasound diagnosis for people. Ultrasound training and EG education were provided at health centers and routine surveillance was conducted in abattoirs. Over five years, dog prevalence dropped from 10% to 1.6%^35^. In Chile’s Aysen region, from 1982 to 2001, dogs were usually dosed every 6 weeks. Human prevalence decreased from 75% to 32% and sheep prevalence from 90% to 9%. In 2020, sheep vaccination was added, and surveillance was enhanced by examining sheep cysts and by communities reporting cysts via WhatsApp. Cyst size was negatively correlated with vaccination status^83^. In another study in Chile, infected dogs were identified and treated with PZQ. Health education was provided to farm family members, and they were asked to pledge to educate their neighbors, to practice what was learned, to buy PZQ as a group for their dogs. Serology was conducted on dog-owners. Unfortunately, only pre-intervention prevalence was reported^84^. In the US, a decade-long control program in Utah involved PZQ treatment for dogs (frequency unspecified), educational initiatives through press releases, filmstrips, and children’s coloring books, diagnostic clinics for humans and dogs, and surveillance and proper disposal of viscera at abattoirs. Over seven years, infection rates in dogs dropped from 28% to 1%, but later rose to 10% due to some dog owners’ noncompliance. Concurrently, EG infection in sheep steadily declined, and public knowledge about the disease significantly increased^85^.

**Table 1c.**
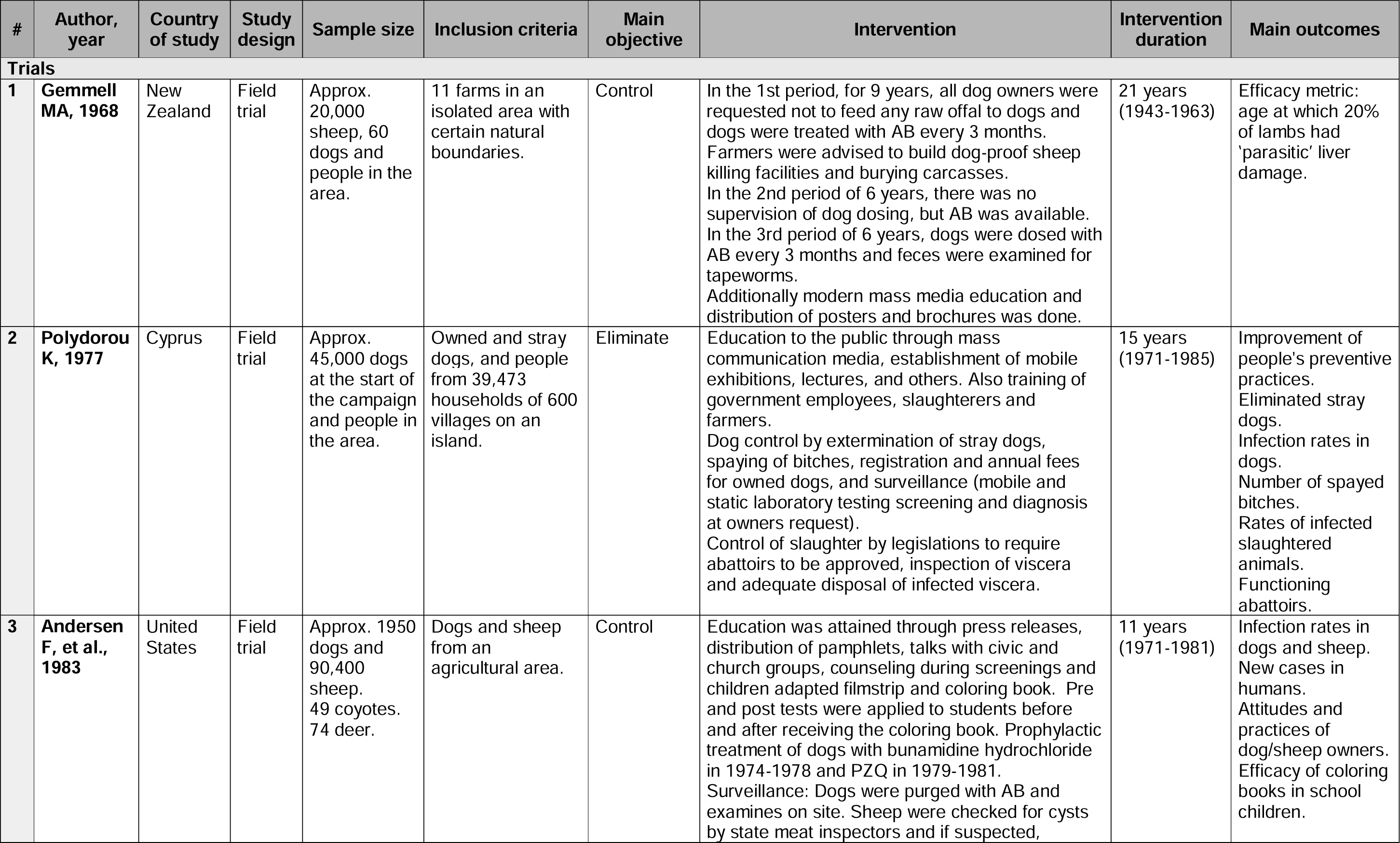
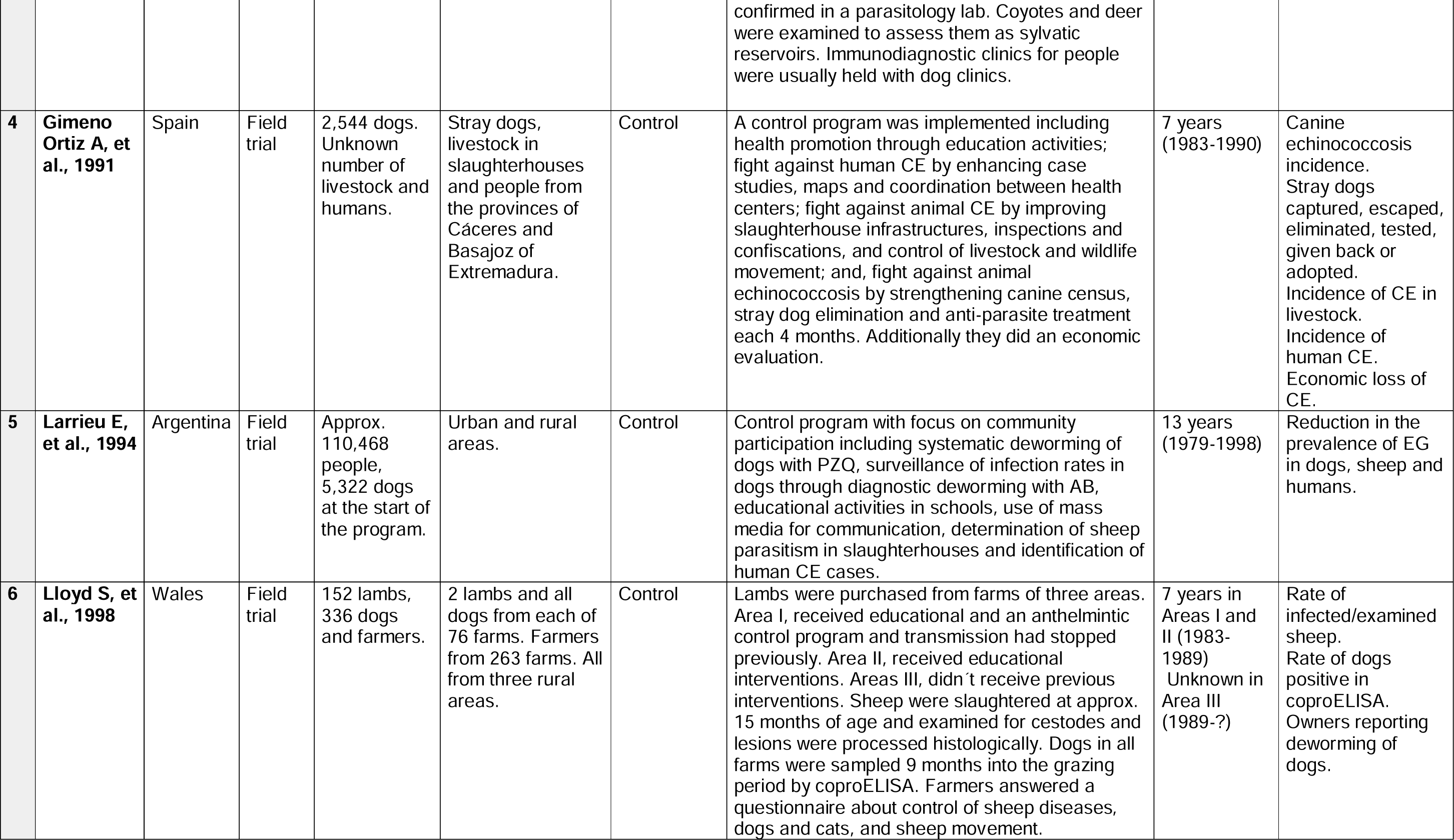
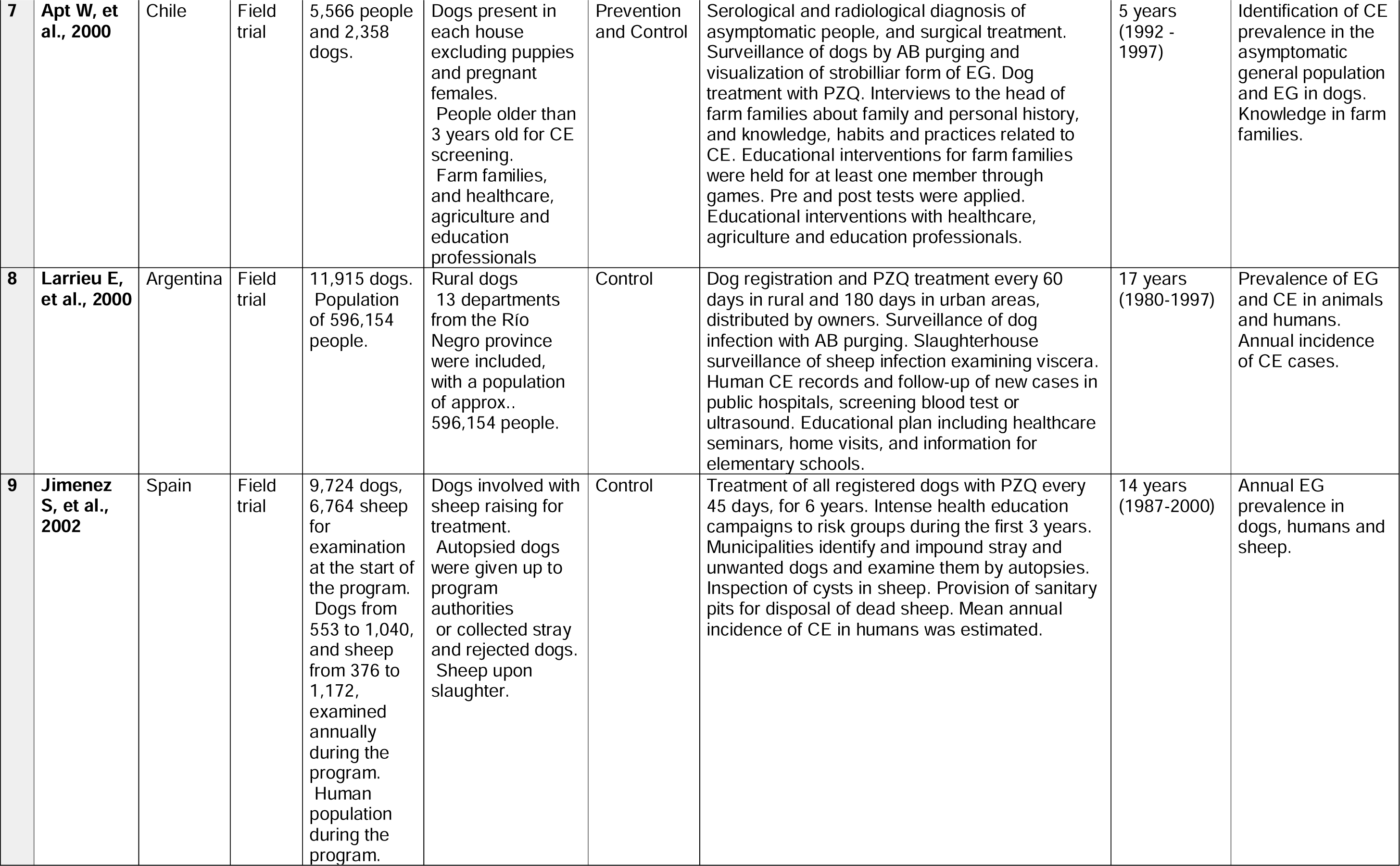
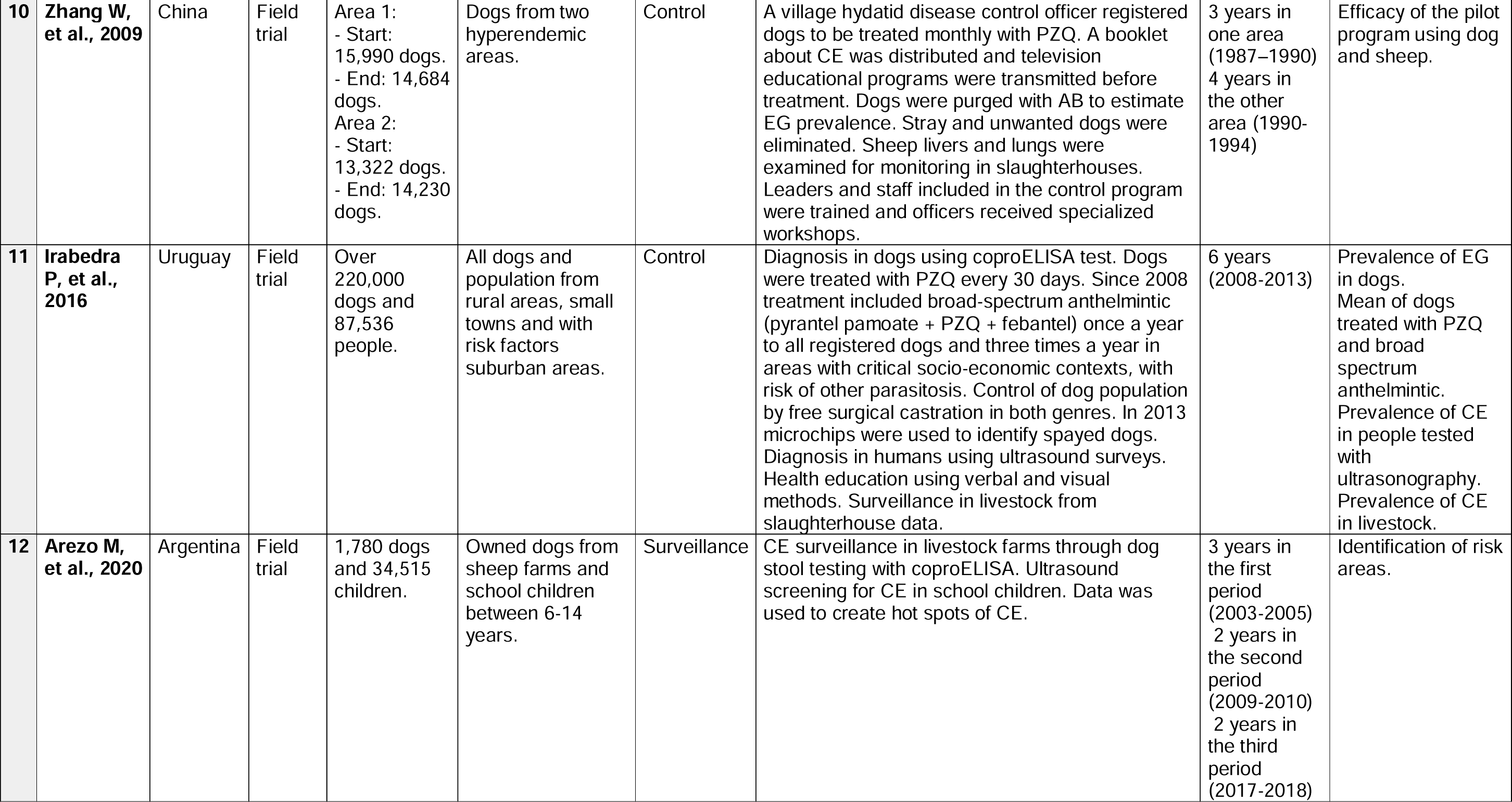
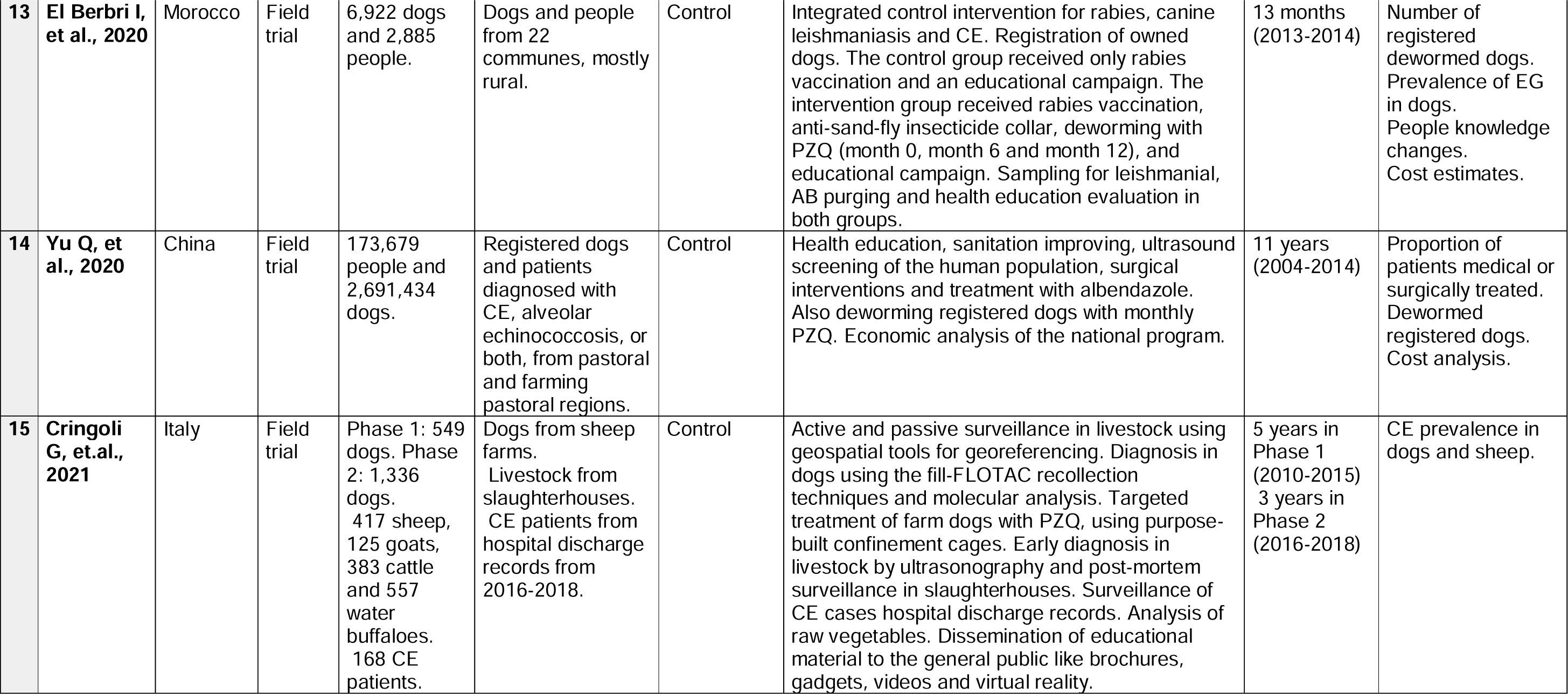
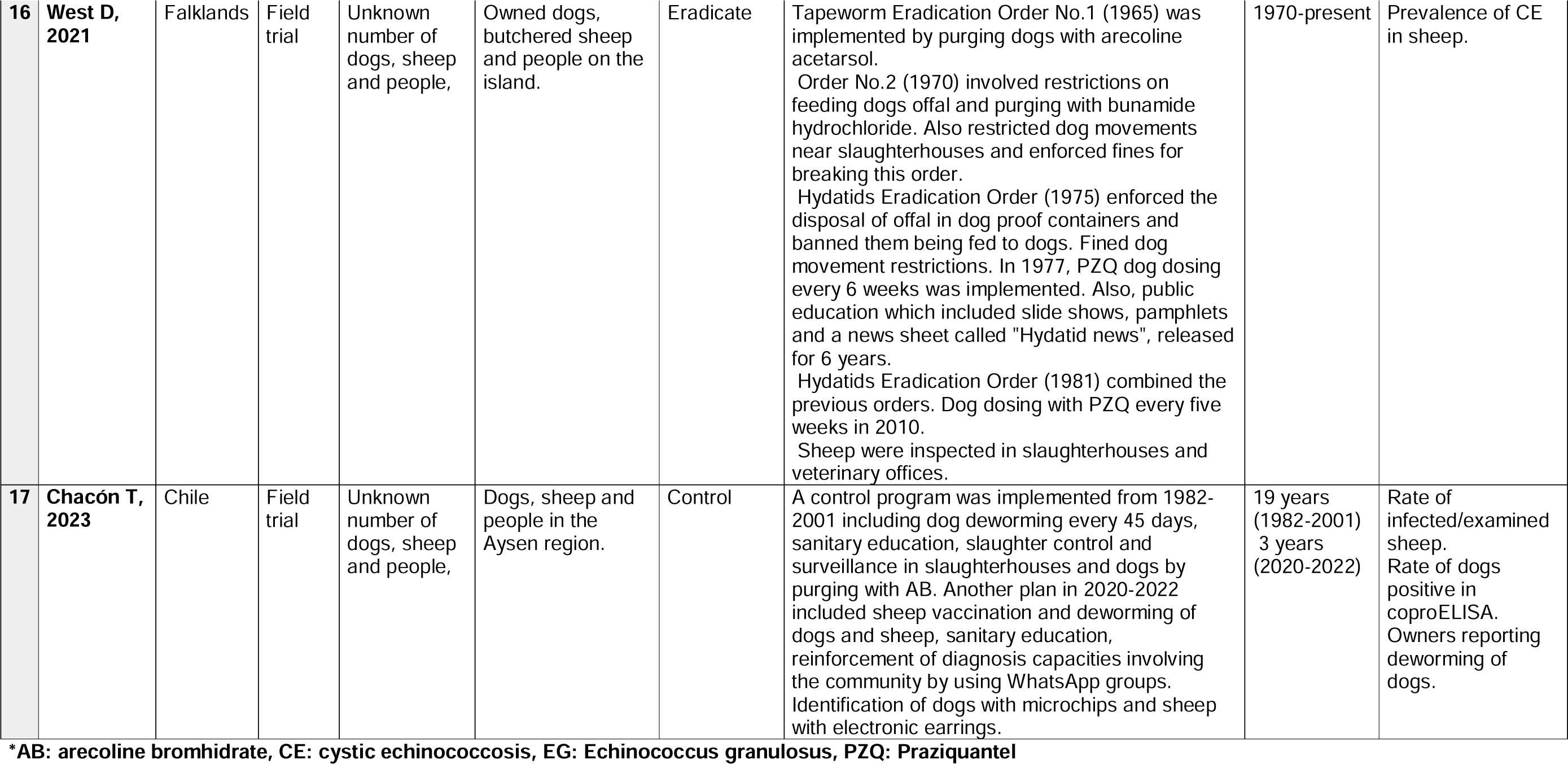
Interventions in Humans and Animals.

| # | Author, year | Country of study | Study design | Sample size | Inclusion criteria | Main objective | Intervention | Intervention duration | Main outcomes |
| --- | --- | --- | --- | --- | --- | --- | --- | --- | --- |
| **Trials** |  |  |  |  |  |  |  |  |  |
| 1 | **Gemmell MA, 1968** | New Zealand | Field trial | Approx. 20,000 sheep, 60 dogs and people in the area. | 11 farms in an isolated area with certain natural boundaries. | Control | In the 1st period, for 9 years, all dog owners were requested not to feed any raw offal to dogs and dogs were treated with AB every 3 months. Farmers were advised to build dog-proof sheep killing facilities and burying carcasses. In the 2nd period of 6 years, there was no supervision of dog dosing, but AB was available. In the 3rd period of 6 years, dogs were dosed with AB every 3 months and feces were examined for tapeworms. Additionally modern mass media education and distribution of posters and brochures was done. | 21 years (1943-1963) | Efficacy metric: age at which 20% of lambs had 'parasitic' liver damage. |
| 2 | **Polydorou K, 1977** | Cyprus | Field trial | Approx. 45,000 dogs at the start of the campaign and people in the area. | Owned and stray dogs, and people from 39,473 households of 600 villages on an island. | Eliminate | Education to the public through mass communication media, establishment of mobile exhibitions, lectures, and others. Also training of government employees, slaughterers and farmers. Dog control by extermination of stray dogs, spaying of bitches, registration and annual fees for owned dogs, and surveillance (mobile and static laboratory testing screening and diagnosis at owners request). Control of slaughter by legislations to require abattoirs to be approved, inspection of viscera and adequate disposal of infected viscera. | 15 years (1971-1985) | Improvement of people's preventive practices. Eliminated stray dogs. Infection rates in dogs. Number of spayed bitches. Rates of infected slaughtered animals. Functioning abattoirs. |
| 3 | **Andersen F, et al., 1983** | United States | Field trial | Approx. 1950 dogs and 90,400 sheep. 49 coyotes. 74 deer. | Dogs and sheep from an agricultural area. | Control | Education was attained through press releases, distribution of pamphlets, talks with civic and church groups, counseling during screenings and children adapted filmstrip and coloring book. Pre and post tests were applied to students before and after receiving the coloring book. Prophylactic treatment of dogs with bunamidine hydrochloride in 1974-1978 and PZQ in 1979-1981. Surveillance: Dogs were purged with AB and examines on site. Sheep were checked for cysts by state meat inspectors and if suspected, confirmed in a parasitology lab. Coyotes and deer were examined to assess them as sylvatic reservoirs. Immunodiagnostic clinics for people were usually held with dog clinics. | 11 years (1971-1981) | Infection rates in dogs and sheep. New cases in humans. Attitudes and practices of dog/sheep owners. Efficacy of coloring books in school children. |
| 4 | **Gimeno Ortiz A, et al., 1991** | Spain | Field trial | 2,544 dogs. Unknown number of livestock and humans. | Stray dogs, livestock in slaughterhouses and people from the provinces of Cáceres and Basajoz of Extremadura. | Control | A control program was implemented including health promotion through education activities; fight against human CE by enhancing case studies, maps and coordination between health centers; fight against animal CE by improving slaughterhouse infrastructures, inspections and confiscations, and control of livestock and wildlife movement; and, fight against animal echinococcosis by strengthening canine census, stray dog elimination and anti-parasite treatment each 4 months. Additionally they did an economic evaluation. | 7 years (1983-1990) | Canine echinococcosis incidence. Stray dogs captured, escaped, eliminated, tested, given back or adopted. Incidence of CE in livestock. Incidence of human CE. Economic loss of CE. |
| 5 | **Larrieu E, et al., 1994** | Argentina | Field trial | Approx. 110,468 people, 5,322 dogs at the start of the program. | Urban and rural areas. | Control | Control program with focus on community participation including systematic deworming of dogs with PZQ, surveillance of infection rates in dogs through diagnostic deworming with AB, educational activities in schools, use of mass media for communication, determination of sheep parasitism in slaughterhouses and identification of human CE cases. | 13 years (1979-1998) | Reduction in the prevalence of EG in dogs, sheep and humans. |
| 6 | **Lloyd S, et al., 1998** | Wales | Field trial | 152 lambs, 336 dogs and farmers. | 2 lambs and all dogs from each of 76 farms. Farmers from 263 farms. All from three rural areas. | Control | Lambs were purchased from farms of three areas. Area I, received educational and an anthelmintic control program and transmission had stopped previously. Area II, received educational interventions. Areas III, didn't receive previous interventions. Sheep were slaughtered at approx. 15 months of age and examined for cestodes and lesions were processed histologically. Dogs in all farms were sampled 9 months into the grazing period by coproELISA. Farmers answered a questionnaire about control of sheep diseases, dogs and cats, and sheep movement. | 7 years in Areas I and II (1983-1989) Unknown in Area III (1989-?) | Rate of infected/examined sheep. Rate of dogs positive in coproELISA. Owners reporting deworming of dogs. |
| 7 | **Apt W, et al., 2000** | Chile | Field trial | 5,566 people and 2,358 dogs. | Dogs present in each house excluding puppies and pregnant females.<br>People older than 3 years old for CE screening.<br>Farm families, and healthcare, agriculture and education professionals | Prevention and Control | Serological and radiological diagnosis of asymptomatic people, and surgical treatment. Surveillance of dogs by AB purging and visualization of strobiliar form of EG. Dog treatment with PZQ. Interviews to the head of farm families about family and personal history, and knowledge, habits and practices related to CE. Educational interventions for farm families were held for at least one member through games. Pre and post tests were applied. Educational interventions with healthcare, agriculture and education professionals. | 5 years (1992 - 1997) | Identification of CE prevalence in the asymptomatic general population and EG in dogs. Knowledge in farm families. |
| 8 | **Larrieu E, et al., 2000** | Argentina | Field trial | 11,915 dogs. Population of 596,154 people. | Rural dogs 13 departments from the Río Negro province were included, with a population of approx. 596,154 people. | Control | Dog registration and PZQ treatment every 60 days in rural and 180 days in urban areas, distributed by owners. Surveillance of dog infection with AB purging. Slaughterhouse surveillance of sheep infection examining viscera. Human CE records and follow-up of new cases in public hospitals, screening blood test or ultrasound. Educational plan including healthcare seminars, home visits, and information for elementary schools. | 17 years (1980-1997) | Prevalence of EG and CE in animals and humans. Annual incidence of CE cases. |
| 9 | **Jimenez S, et al., 2002** | Spain | Field trial | 9,724 dogs, 6,764 sheep for examination at the start of the program. Dogs from 553 to 1,040, and sheep from 376 to 1,172, examined annually during the program. Human population during the program. | Dogs involved with sheep raising for treatment. Autopsied dogs were given up to program authorities or collected stray and rejected dogs. Sheep upon slaughter. | Control | Treatment of all registered dogs with PZQ every 45 days, for 6 years. Intense health education campaigns to risk groups during the first 3 years. Municipalities identify and impound stray and unwanted dogs and examine them by autopsies. Inspection of cysts in sheep. Provision of sanitary pits for disposal of dead sheep. Mean annual incidence of CE in humans was estimated. | 14 years (1987-2000) | Annual EG prevalence in dogs, humans and sheep. |
| 10 | **Zhang W, et al., 2009** | China | Field trial | Area 1:<br>- Start: 15,990 dogs.<br>- End: 14,684 dogs.<br>Area 2:<br>- Start: 13,322 dogs.<br>- End: 14,230 dogs. | Dogs from two hyperendemic areas. | Control | A village hydatid disease control officer registered dogs to be treated monthly with PZQ. A booklet about CE was distributed and television educational programs were transmitted before treatment. Dogs were purged with AB to estimate EG prevalence. Stray and unwanted dogs were eliminated. Sheep livers and lungs were examined for monitoring in slaughterhouses. Leaders and staff included in the control program were trained and officers received specialized workshops. | 3 years in one area (1987–1990)<br>4 years in the other area (1990–1994) | Efficacy of the pilot program using dog and sheep. |
| 11 | **Irabedra P, et al., 2016** | Uruguay | Field trial | Over 220,000 dogs and 87,536 people. | All dogs and population from rural areas, small towns and with risk factors suburban areas. | Control | Diagnosis in dogs using coproELISA test. Dogs were treated with PZQ every 30 days. Since 2008 treatment included broad-spectrum anthelmintic (pyrantel pamoate + PZQ + febantel) once a year to all registered dogs and three times a year in areas with critical socio-economic contexts, with risk of other parasitosis. Control of dog population by free surgical castration in both genres. In 2013 microchips were used to identify spayed dogs. Diagnosis in humans using ultrasound surveys. Health education using verbal and visual methods. Surveillance in livestock from slaughterhouse data. | 6 years (2008-2013) | Prevalence of EG in dogs. Mean of dogs treated with PZQ and broad spectrum anthelmintic. Prevalence of CE in people tested with ultrasonography. Prevalence of CE in livestock. |
| 12 | **Arezo M, et al., 2020** | Argentina | Field trial | 1,780 dogs and 34,515 children. | Owned dogs from sheep farms and school children between 6-14 years. | Surveillance | CE surveillance in livestock farms through dog stool testing with coproELISA. Ultrasound screening for CE in school children. Data was used to create hot spots of CE. | 3 years in the first period (2003-2005)<br>2 years in the second period (2009-2010)<br>2 years in the third period (2017-2018) | Identification of risk areas. |
| 13 | **El Berbri I, et al., 2020** | Morocco | Field trial | 6,922 dogs and 2,885 people. | Dogs and people from 22 communes, mostly rural. | Control | Integrated control intervention for rabies, canine leishmaniasis and CE. Registration of owned dogs. The control group received only rabies vaccination and an educational campaign. The intervention group received rabies vaccination, anti-sand-fly insecticide collar, deworming with PZQ (month 0, month 6 and month 12), and educational campaign. Sampling for leishmanial, AB purging and health education evaluation in both groups. | 13 months (2013-2014) | Number of registered dewormed dogs. Prevalence of EG in dogs. People knowledge changes. Cost estimates. |
| 14 | **Yu Q, et al., 2020** | China | Field trial | 173,679 people and 2,691,434 dogs. | Registered dogs and patients diagnosed with CE, alveolar echinococcosis, or both, from pastoral and farming pastoral regions. | Control | Health education, sanitation improving, ultrasound screening of the human population, surgical interventions and treatment with albendazole. Also deworming registered dogs with monthly PZQ. Economic analysis of the national program. | 11 years (2004-2014) | Proportion of patients medical or surgically treated. Dewormed registered dogs. Cost analysis. |
| 15 | **Cringoli G, et.al., 2021** | Italy | Field trial | Phase 1: 549 dogs. Phase 2: 1,336 dogs. 417 sheep, 125 goats, 383 cattle and 557 water buffaloes. 168 CE patients. | Dogs from sheep farms. Livestock from slaughterhouses. CE patients from hospital discharge records from 2016-2018. | Control | Active and passive surveillance in livestock using geospatial tools for georeferencing. Diagnosis in dogs using the fill-FLOTAC recollection techniques and molecular analysis. Targeted treatment of farm dogs with PZQ, using purpose-built confinement cages. Early diagnosis in livestock by ultrasonography and post-mortem surveillance in slaughterhouses. Surveillance of CE cases hospital discharge records. Analysis of raw vegetables. Dissemination of educational material to the general public like brochures, gadgets, videos and virtual reality. | 5 years in Phase 1 (2010-2015)<br>3 years in Phase 2 (2016-2018) | CE prevalence in dogs and sheep. |
| 16 | West D, 2021 | Falklands | Field trial | Unknown number of dogs, sheep and people, | Owned dogs, butchered sheep and people on the island. | Eradicate | Tapeworm Eradication Order No.1 (1965) was implemented by purging dogs with arecoline acetarsol.<br>Order No.2 (1970) involved restrictions on feeding dogs offal and purging with bunamide hydrochloride. Also restricted dog movements near slaughterhouses and enforced fines for breaking this order.<br>Hydatids Eradication Order (1975) enforced the disposal of offal in dog proof containers and banned them being fed to dogs. Fined dog movement restrictions. In 1977, PZQ dog dosing every 6 weeks was implemented. Also, public education which included slide shows, pamphlets and a news sheet called "Hydatid news", released for 6 years.<br>Hydatids Eradication Order (1981) combined the previous orders. Dog dosing with PZQ every five weeks in 2010.<br>Sheep were inspected in slaughterhouses and veterinary offices. | 1970-present | Prevalence of CE in sheep. |
| 17 | Chacón T, 2023 | Chile | Field trial | Unknown number of dogs, sheep and people, | Dogs, sheep and people in the Aysen region. | Control | A control program was implemented from 1982-2001 including dog deworming every 45 days, sanitary education, slaughter control and surveillance in slaughterhouses and dogs by purging with AB. Another plan in 2020-2022 included sheep vaccination and deworming of dogs and sheep, sanitary education, reinforcement of diagnosis capacities involving the community by using WhatsApp groups. Identification of dogs with microchips and sheep with electronic earrings. | 19 years (1982-2001)<br>3 years (2020-2022) | Rate of infected/examined sheep.<br>Rate of dogs positive in coproELISA.<br>Owners reporting deworming of dogs. |

\*AB: arecoline bromhidrate, CE: cystic echinococcosis, EG: Echinococcus granulosus, PZQ: Praziquantel

Studies and programs from other parts of the world used a similar integrative approach. In New Zealand, a 21-year program involved treating dogs regularly with arecoline, banning raw offal feeding, and promoting safe sheep slaughtering practices. The authors used the age at which 20% of lambs had ‘parasitic’ liver damage as an efficacy metric. Over the trial, this age increased from 4 to 9 months^86^. In Cyprus, education was carried out over 5 years, targeting school children, housewives, and dog owners. Dogs were screened and infected dogs were euthanized or arecoline was given according to the owner’s consent. After three years, dog prevalence dropped from 50% to 6.8%^87^. In Wales and Morocco education alone was compared to education plus PZQ. In Wales, after the intervention, lamb prevalence was 4.3% in the education area, 6% in the education plus PZQ area, and 10.4% in the control area. Education significantly increased PZQ usage among farmers^88^. In Morocco, an integrated intervention targeted dog rabies, echinococcosis, and canine leishmaniasis. Dogs were treated 3 times a year and purged with arecoline at the end of the study. Dogs receiving PZQ had lower infection rates than those in the only education arm, but the difference was not statistically significant. Education increased knowledge but did not affect risky behaviors^89^. In Spain, dogs were treated with PZQ every 6 weeks for 6 years, health education targeted high-risk groups for the first 3 years, and stray dogs were removed. Sheep were inspected and sanitary pits were provided to dispose of carcasses. New human cases decreased from 19 to 4 per 100,000 people^90^. Also in Spain, a comprehensive program is reported though it lacks detailed descriptions of each component. Nonetheless, the program achieved an overall efficacy of 67% and, importantly, reported a profitability of 970%^91^. In China, in hyperendemic areas a combination of monthly PZQ, culling unwanted dogs, restructuring and training of the EG management teams, and public education reduced prevalence in dogs from 14.7-18.6% to zero after 4 years^92^. Another intervention consisted of education, improved sanitation, ultrasound screening of humans, surgical and albendazole treatment, and management and monthly PZQ for dogs. Interestly, the authors report measures of efficacy different from disease burden. After 11 years, surgeries rose by 32.4%, albendazole treatment by 81.3%, and dog deworming by 58.6%^93^. In Italy, an 8-year program used sentinel sheep abattoirs, analyzed national slaughterhouse data annually, treated farm dogs with PZQ, and diagnosed dogs with molecular and fill-FLOTAC techniques. Livestock were diagnosed with ultrasound and human CE surveillance was based on medical records. Public health education was delivered through multiple media formats. The program reduced sheep prevalence by 30%, and all dogs were negative 50 days post-PZQ^44^. In the Falklands, legally imposed control activities between 1965 and 1981 included purging dogs, banning offal feeding, and enforcing shepherd dog movement restrictions. Dogs were treated with PZQ every 6 weeks. Education was conducted through various media formats. Sheep prevalence dropped from over 40% to below 1% by 1993^94^. Details of these interventions focused on animals and humans can be found in Table 1c.

In Fig 2, we illustrate the points within the EG life cycle where different studies aimed to interrupt transmission. As numerous studies implemented multiple interventions targeting various components of the life cycle, the count of interventions depicted in Fig 2 exceeds the total number of papers analyzed.

### Quality Appraisal

Of the 45 studies in our review, 40 were evaluated with the quantitative non-randomized studies questions (section 3 of MMAT). The average fulfillment with the quality criteria was 57%: five studies^59,70,83,86,91^ did not meet any criteria or their compliance could not be determined, three^58,71,88^ met 20%, one^84^ met 40%, 16 studies^44,54,61,62,65,72,73,75,78,80,81,87,89,90,93,94^ met 60%, 14 studies^35,55,61,63,64,66,67,74,76,77,79,82,85,92^ met 80%, and only one study^68^ met 100% of the quality criteria. The main criterion for noncompliance/undetermined compliance was the evaluation of possible confounding factors; only one study^68^ fully addressed them. The second most frequent challenge was determining representativeness; sampling estimates or clear description of sample selection were not always provided. Another criterion with inadequate or undetermined answers was completeness of the outcome data. Three studies were evaluated with the quantitative descriptive study questions (section 4 of MMAT). The average overall quality was 47%; one^39^ met 20% of the criteria, one^53^ met 40%, and one^56^ met 80%. The main criterion for compliance was the evaluation of risk of nonresponse bias and we found that the information provided was insufficient to comply with the criterion. The second most frequent problem was evaluation of representativeness. The three studies described their target population and eligibility, but in two^39,53^ it was difficult to determine if their sample was representative. One study^69^ was evaluated with the quantitative randomized controlled trials questions (section 2 of MMAT), and it met 60% of the quality criteria. In this study, the information provided was insufficient to fully evaluate if the randomization was appropriately performed and no information about blinding was provided. One study^57^ was evaluated with the mixed methods study questions (sections 5, 1 and 4 of MMAT) and met 40% of the overall quality criteria. This mixed methods study had a predominance of qualitative methods and results. When evaluated by components, it scored 100% of compliance in the qualitative criteria (section 1), 60% in the mixed methods criteria (section 5) and 40% in the quantitative criteria (section 4).

## Discussion

We mapped and analyzed 45 articles on interventions for the control, prevention, or elimination of EG from all regions (Fig 3). The primary goal of control interventions is to halt the transmission of EG to its hosts. Compared to earlier interventions^72,86,87^, recent advances have significantly enhanced EG control. Key developments include the incorporation of the EG95 vaccine for sheep^95^, improved diagnostic techniques for early detection in dogs (e.g., ELISA)^96,97^, and the use of PZQ for dog treatment^98^. Despite these advances, progress in EG control remains inconsistent, with mixed results reported^99^, and overall, limited advancement in endemic regions^100–102^. We found heterogeneous and generally low quality articles, often lacking clear post-intervention outcomes, which hampers the ability to inform future interventions. Studies varied in scale but provided limited information for scale-up implementation or replication in different settings. Common barriers are often mentioned, but few studies explore real-world barriers and facilitators in depth or address the feasibility of interventions. Additionally, implementation costs are rarely reported, yet cost-benefit analyses are essential for the scalability and sustainability of these strategies^103–105^. In addition, effective EG control requires collaborations, partnerships, and community engagement. The educational interventions we reviewed here were aimed at increasing awareness and knowledge among various stakeholders^39,54–57^, but did not report measures of long-term impact. However, the primary value of these interventions may lie in promoting community engagement, which is crucial for the success of control and elimination efforts^106,107^. Future studies could aim at building and examining stakeholder and community engagement for enforcement and sustainability of control activities.

Among the reviewed studies, those focused solely on interventions on animals were the most numerous, while studies focused solely on interventions on humans were the fewest. However, studies focused on both animals and humans had, in general, more participants, lasted longer and covered larger geographical areas. The majority of these articles on integrated strategies described national multi-pronged programs including population screening and diagnosis, sanitation improvement, human and animal treatment and surveillance, among others. However, most of these articles came from a few countries who have been able to sustain these massive efforts to control EG. However, a significant challenge in the control programs for cystic echinococcosis (CE) is ensuring government commitment^108,109^. These programs require substantial effort and financial investment, funding that should come from government sources, such as Ministries of Health and Ministries of Agriculture, and participation of other sectors such as education and environment^109,110^. Short-term programs spanning only a few years are inadequate; CE must be treated as a chronic endemic disease requiring sustained efforts over many years^18,111^. Countries, such as Uruguay and Argentina, have been implementing control programs for over 50 years with important successes but they continue to face challenges despite their long-term commitment^30,112^.

Others have conducted reviews on EG and its control. For ours, we used the PRISMA-ScR framework and the Arksey and O’Malley method, focusing on studies in English, Spanish, and Portuguese. Others have used various guidelines and checklists, including PRISMA^15,113^, MeSH^26^, and Medline^18^, to evaluate articles across multiple languages and databases. We used MMAT for quality appraisal, while others used RCVS, CASP and JBI^114^. Despite the lack of detailed methodologies in other reviews on EG, they provide comprehensive summaries of the determinants and control programs associated with EG infection. A review^26^ highlighted projects like HERACLES (Human Echinococcosis ReseArch in Central and Eastern Societies)^115,116^ and MEmE (Multi-centre study on *Echinococcus multilocularis* and EG in Europe: development and harmonization of diagnostic methods in the food chain)^117,118^, which have created a comprehensive database with the latest epidemiological data from various regions. Multiple reviews^18,26,119,120^ agreed with us that sheep vaccination emerged as a promising strategy to complement and reduce the time of the control efforts. However, it is important to recognize that logistical and financial challenges remain in endemic countries, impacting the effectiveness of these interventions.

Similar to our results^87,94^, others have found that island control programs tend to have higher success rates possibly due to easier disease containment and more manageable populations^99^. Interestingly, one review suggested sustained vertical campaigns that include treatment of dogs, community education, surveillance in rural areas, although they warn about significant time and resources^99^. We found vertical programs with strict enforcement measures in the Falklands and Cyprus^87,94^, both with positive results; however, the insular context might have played a bigger role in the success of those programs. Key challenges reported in these reviews and also found in our review are reliance on rural dog owners for implementing dog treatment, inadequate funding and staffing of programs, and political instability or insufficient political will^6,18,119^. The One-Health approach is recommended to address these issues effectively^120–122^. Other frequently encountered implementation barriers that most studies fail to report are limited surveillance^71,81^, followed by the presence of ownerless dogs^60,81,89^, and two barriers that are assumed common in many endemic countries: geographic accessibility and the semi-nomadic lifestyle of some communities^73^.

Multiple animal-targeting studies reviewed here have tested and refined interventions for the main definitive EG host, the domestic dogs. Since the development of PZQ (Fig 4), an effective cestocide for the mature and immature adult stages of EG^123^, that drug has been the primary intervention for dogs, with treatment schemes varying from monthly to quarterly, depending on the disease burden in the area. Challenges for the effectiveness of PZQ include maintaining treatment, achieving high coverage, owner compliance, and government enforcement^18,124^. One of the fundamental causes for not achieving high coverage is the inability to find the dog at the house to receive the treatment. Recent innovative interventions involve advanced tools like geolocation devices to treat hotspots, smart collars for automatic baiting, and dropping PZQ baits with drones^74,77,79^, which could improve efficiency and reduce costs in control campaigns. Currently, logistical costs are too high for sustained PZQ-based control programs^125,126^. Some of the studies we discuss here report a payment from the dog owners^127^, others promote dog owners to buy PZQ in bulk to treat their animals^84^, other require an annual registration fee for the dog to support PZQ treatment among other dog-related interventions^87^, while in a few countries PZQ is provide free of cost to dog owners^63,73,81^.

**Figure 4.**
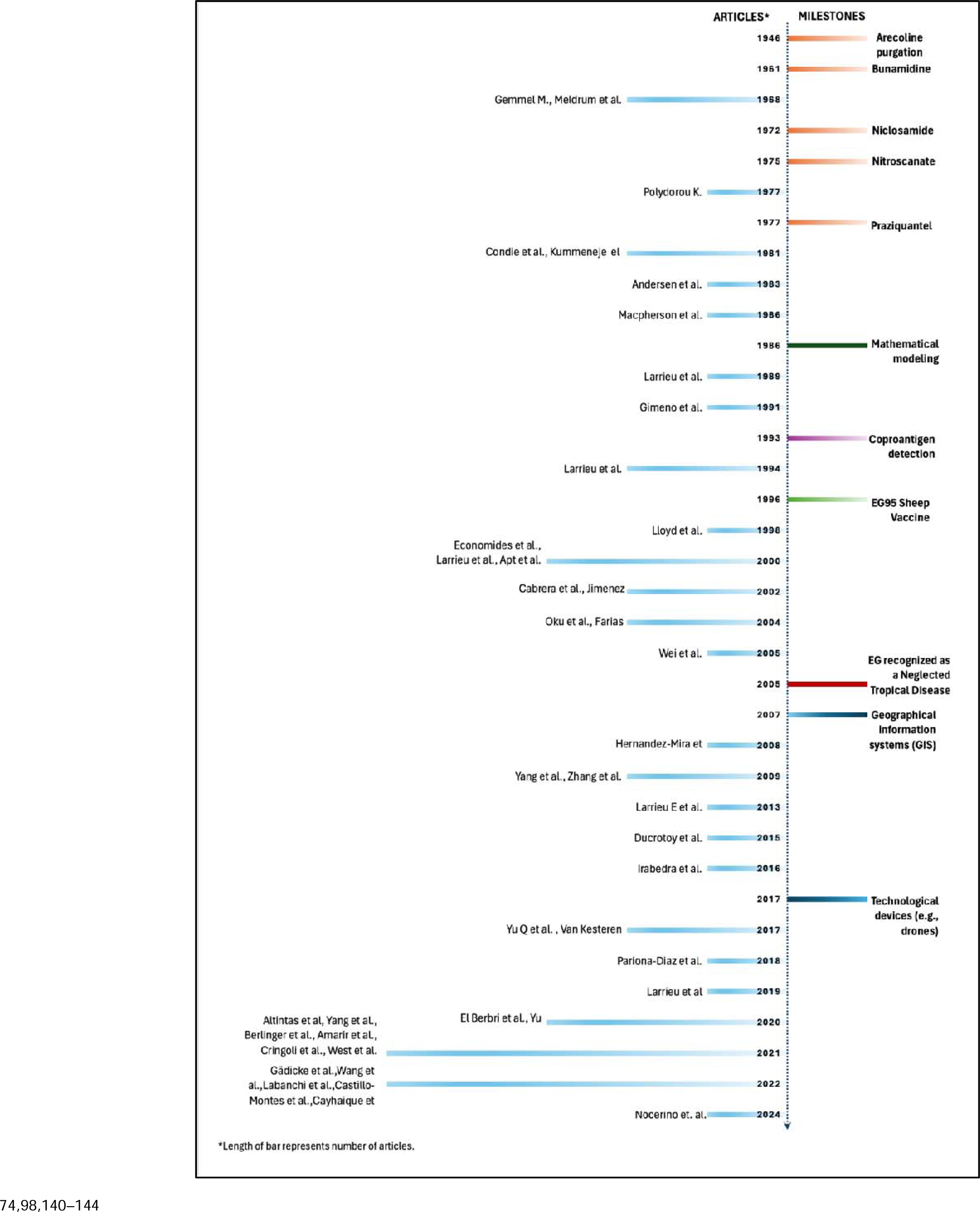
Timeline of the progress of EG control tools and the articles included in this review.

From the beginning of PZQ and other oral treatments for dogs, palatability and acceptability studies have been conducted to improve mass use of these drugs^128^. More recent studies have focused on understanding reinfection rates after PZQ^125^ and testing slow-release PZQ formulations to reduce frequency of treatment^76,126^. A formulation that protects dogs for long periods (similar to what is expected from a vaccine) would be a real game changer. Equally important are studies on stakeholder acceptability of interventions. For example, some studies found conflicting perspectives coming from breeders and their wifes, and local authorities, and health providers related to EG control actions^38,129^. To enhance the feasibility, sustainability, and ultimate success of elimination and control programs, it is imperative to conduct implementation research^130,131^ studies that delve into the agendas and motivations of various stakeholders^132,133^. Understanding the perspectives and priorities of these stakeholders can increase community engagement and inform the design and implementation of more effective interventions^133^. Active participation from the community ensures better acceptance and adherence to control measures, fostering a sense of ownership and responsibility towards the program’s goals^134,135^. This multifaceted program would align well with the One Health approach^136–138^ not only addressing the technical and logistical challenges, but also building a supportive environment that can sustain long-term efforts against the disease.

Our study had some limitations. The final set of articles included a wide range of study designs and approaches, which led to significant heterogeneity in the data. This heterogeneity made it challenging to summarize findings across studies. Differences in the reported methods and results between studies made the classification of studies especially challenging for the quality appraisal. As with other types of reviews, our scoping review could be susceptible to publication bias, due to unpublished or inaccessible studies. Also, we made a strong effort to balance breadth and depth; however, it is possible we prioritized more breadth making it difficult to synthesize findings comprehensively. A challenge we faced was the reduced number of studies included for full-text review. With sparse data, generalizability may be limited and there may be a risk of over interpreting the significance of findings. Moreover, this limited number of studies impeded identifying trends. Despite these limitations, we conducted a rigorous quality assessment of the included studies and implemented a detailed protocol to reduce subjectivity in study selection and data extraction.

## Conclusion

In sum, the literature on control, prevention, or elimination of *E. granulosus sensu lato* shows key advancements in diagnosis, treatment, and vaccines for the intermediate and definitive hosts. However, progress remains inconsistent, particularly in endemic regions, and many studies lack clear post-intervention outcomes, impeding the ability to inform future programs. Standardization in the reporting of studies is necessary for comparability of results and scaling-up of interventions. Effective EG control requires sustained government commitment, funding, and active participation from various sectors, including health, agriculture, and education. Long-term programs are necessary, as demonstrated by the decades-long efforts in Uruguay and Argentina. Innovative strategies, such as geolocation devices and drones for PZQ baiting, show promise but face logistical and financial challenges. To enhance the feasibility and sustainability of elimination and control programs, implementation research focused on understanding stakeholders’ agendas and increasing community engagement is crucial. Multi-pronged integrative One Health approaches are essential for overcoming the logistical, economical, and political challenges of sustaining long-term efforts for echinococcosis control.

## Data Availability

The protocol of this review is registered on the Open Science Framework (www.osf.io) with DOI 10.17605/OSF.IO/48AZR. URL: https://osf.io/agsfh URL: https://osf.io/48azr All other articles included in results of this scoping review are published in their respective journals.

## Acknowledgments

We are grateful to Fredy Daniel Bustamante Villanueva for his support during article retrieval.

## Supporting information

### Appendix 1. Search Strategy

Search Concepts

|  |  |
| --- | --- |
| Echinococcus Granulosus | Strategies |
| Echinococcus | Prevention |
| Echinococcosis | Control |
| Echinococcoses | Preventive therapy |
| Echinococcus Infection | Preventive measures |
| Echinococcus Infections | Prophylaxis |
| Infection, Echinococcus | Therapeutic |
| Cystic Echinococcosis | Therapy |
| Cystic Echinococcoses | Treatment |
| Hydatidosis | Treatments |
| Hydatidoses | Elimination |
| Hydatid Cysts | Intervention |
| Hydatid Cyst | Early intervention |
| Hydatid Disease | Late intervention |
| Hydatid Diseases | Drug therapy |
| Echinococcus Granulosus Infection | Drug Therapies |
|  | Pharmacotherapy |
|  | Pharmacotherapies |
|  | Vaccine |
|  | Health Promotion |
|  | Health Campaign |
|  | Pest control |
|  | Pest management |

MEDLINE Search strategy:

> 1. Echinococcus Granulosus
>
> 2. Echinococcus
>
> 3. Echinococcosis
>
> 4. Echinococcoses
>
> 5. Echinococcus Infection
>
> 6. Echinococcus Infections
>
> 7. Infection, Echinococcus
>
> 8. Cystic Echinococcosis
>
> 9. Cystic Echinococcoses
>
> 10. Hydatidosis
>
> 11. Hydatidoses
>
> 12. Hydatid Cysts
>
> 13. Hydatid Cyst
>
> 14. Hydatid Disease
>
> 15. Hydatid Diseases
>
> 16. Echinococcus Granulosus Infection
>
> 17. 1-16 OR
>
> 18. Strategies
>
> 19. Prevention
>
> 20. Control
>
> 21. Preventive therapy
>
> 22. Preventive measures
>
> 23. Prophylaxis
>
> 24. Therapeutic
>
> 25. Therapy
>
> 26. Treatment
>
> 27. Treatments
>
> 28. Elimination
>
> 29. Intervention
>
> 30. Early intervention
>
> 31. Late intervention
>
> 32. Drug therapy
>
> 33. Drug Therapies
>
> 34. Pharmacotherapy
>
> 35. Pharmacotherapies
>
> 36. Vaccine
>
> 37. Health Promotion
>
> 38. Health Campaign
>
> 39. Pest control
>
> 40. Pest management
>
> 41. 18-40 OR
>
> 43. 17 AND 41

For MEDLINE and EMBASE we used MeSH subject heading.

Search terms used are centered on the concepts of ‘*Echinococcus granulosus* infection’ and ‘strategy’. The ‘*Echinococcus granulosus*’ concept was built around synonyms (e.g., Hydatid disease, Hydatid cyst). The “strategy” concept was broad and included terms covering all types of interventions (e.g., Pest control, Therapy, Prevention). Qualified academic librarian support was requested for identification of key words on the different databases and search strategy refinement.

## Appendix 2. Preferred Reporting Items for Systematic reviews and Meta-Analyses extension for Scoping Reviews (PRISMA-ScR) Checklist

| SECTION | ITEM | PRISMA-ScR CHECKLIST ITEM | REPORTED ON PAGE # |
| --- | --- | --- | --- |
| **TITLE** |  |  |  |
| Title | 1 | Identify the report as a scoping review. | 1 |
| **ABSTRACT** |  |  |  |
| Structured summary | 2 | Provide a structured summary that includes (as applicable): background, objectives, eligibility criteria, sources of evidence, charting methods, results, and conclusions that relate to the review questions and objectives. | 2 |
| **INTRODUCTION** |  |  |  |
| Rationale | 3 | Describe the rationale for the review in the context of what is already known. Explain why the review questions/objectives lend themselves to a scoping review approach. | 3-4 |
| Objectives | 4 | Provide an explicit statement of the questions and objectives being addressed with reference to their key elements (e.g., population or participants, concepts, and context) or other relevant key elements used to conceptualize the review questions and/or objectives. | 5-6 |
| **METHODS** |  |  |  |
| Protocol and registration | 5 | Indicate whether a review protocol exists; state if and where it can be accessed (e.g., a Web address); and if available, provide registration information, including the registration number. | 5 |
| Eligibility criteria | 6 | Specify characteristics of the sources of evidence used as eligibility criteria (e.g., years considered, language, and publication status), and provide a rationale. | 6-7, 57 |
| Information sources* | 7 | Describe all information sources in the search (e.g., databases with dates of coverage and contact with authors to identify additional sources), as well as the date the most recent search was executed. | 6 |
| Search | 8 | Present the full electronic search strategy for at least 1 database, including any limits used, such that it could be repeated. | 54-56 |
| Selection of sources of evidence† | 9 | State the process for selecting sources of evidence (i.e., screening and eligibility) included in the scoping review. | 6-7 |
| Data charting process‡ | 10 | Describe the methods of charting data from the included sources of evidence (e.g., calibrated forms or forms that have been tested by the team before their use, and whether data charting was done independently or in duplicate) and any processes for obtaining and confirming data from investigators. | 7 |
| Data items | 11 | List and define all variables for which data were sought and any assumptions and simplifications made. | 7 |
| Critical appraisal of individual sources of evidence§ | 12 | If done, provide a rationale for conducting a critical appraisal of included sources of evidence; describe the methods used and how this information was used in any data synthesis (if appropriate). | 7-8 |
| Synthesis of results | 13 | Describe the methods of handling and summarizing the data that were charted. | 7 |
| **RESULTS** |  |  |  |
| Selection of sources of evidence | 14 | Give numbers of sources of evidence screened, assessed for eligibility, and included in the review, with reasons for exclusions at each stage, ideally using a flow diagram. | 8, 57 |
| Characteristics of sources of evidence | 15 | For each source of evidence, present characteristics for which data were charted and provide the citations. | 8 |
| Critical appraisal within sources of evidence | 16 | If done, present data on critical appraisal of included sources of evidence (see item 12). | 31 |
| Results of individual sources of evidence | 17 | For each included source of evidence, present the relevant data that were charted that relate to the review questions and objectives. | 9, 13-15, 22-24 |
| Synthesis of results | 18 | Summarize and/or present the charting results as they relate to the review questions and objectives. | 10-12, 16-21, 25-30 |
| **DISCUSSION** |  |  |  |
| Summary of evidence | 19 | Summarize the main results (including an overview of concepts, themes, and types of evidence available), link to the review questions and objectives, and consider the relevance to key groups. | 32-35 |
| Limitations | 20 | Discuss the limitations of the scoping review process. | 35-36 |
| Conclusions | 21 | Provide a general interpretation of the results with respect to the review questions and objectives, as well as potential implications and/or next steps. | 36 |
| **FUNDING** |  |  |  |
| Funding | 22 | Describe sources of funding for the included sources of evidence, as well as sources of funding for the scoping review. Describe the role of the funders of the scoping review. | 3 |

From: Tricco AC, Lillie E, Zarin W, O'Brien KK, Colquhoun H, Levac D, et al. PRISMA Extension for Scoping Reviews (PRISMA-ScR): Checklist and Explanation. Ann Intern Med. 2018;169:467–473. doi: [10.7326/M18-0850](https://doi.org/10.7326/M18-0850).

